# Deep neural networks learn general and clinically relevant representations of the ageing brain

**DOI:** 10.1101/2021.10.29.21265645

**Authors:** Esten H. Leonardsen, Han Peng, Tobias Kaufmann, Ingrid Agartz, Ole A. Andreassen, Elisabeth Gulowsen Celius, Thomas Espeseth, Hanne F. Harbo, Einar A. Høgestøl, Ann-Marie de Lange, Andre F. Marquand, Didac Vidal-Piñeiro, James M. Roe, Geir Selbæk, Øystein Sørensen, Stephen M. Smith, Lars T. Westlye, Thomas Wolfers, Yunpeng Wang

## Abstract

The discrepancy between chronological age and the apparent age of the brain based on neuroimaging data — the brain age delta — has emerged as a reliable marker of brain health. With an increasing wealth of data, approaches to tackle heterogeneity in data acquisition are vital. To this end, we compiled raw structural magnetic resonance images into one of the largest and most diverse datasets assembled (n=53542), and trained convolutional neural networks (CNNs) to predict age. We achieved state-of-the-art performance on unseen data from unknown scanners (n=2553), and showed that higher brain age delta is associated with diabetes, alcohol intake and smoking. Using transfer learning, the intermediate representations learned by our model complemented and partly outperformed brain age delta in predicting common brain disorders. Our work shows we can achieve generalizable and biologically plausible brain age predictions using CNNs trained on heterogeneous datasets, and transfer them to clinical use cases.

## Introduction

Neurodevelopmental and age-related changes in the brain play a crucial role in the etiology of complex neurological^1^ and mental disorders^2^. Predictive models for an individuals age based on magnetic resonance imaging (MRI) have been used to estimate normative trajectories across the lifespan^3–6^. Individual deviations from these trajectories, often called the brain age delta, have been linked to brain health^7,8^, and more extreme deviations observed in patients with schizophrenia (SCZ)^9–11^, depression^12^, cognitive impairment^13,14^, dementia^15^, Alzheimer’s disease (AD)^16^ and multiple sclerosis (MS)^17^, implying that such deviations could be a feasible biological marker for various brain disorders.

The brain age of an individual is typically estimated from brain images using statistical learning techniques. The first-generation models were relatively simple, typically based on independent voxels^3^ or a limited number of imaging-derived phenotypes (IDPs) reflecting brain properties such as volumetric measures of different regions^18^. These models generally estimated linear relationships, were restricted in scale, and were trained on datasets with tens, hundreds or a few thousand participants^7^. In parallel with continuous computational advances the exponential growth of MRI data has enabled deep learning models of scale for accurate brain age estimation^6,19^. Deep learning models can take minimally- or non-preprocessed 3-D images as input - avoiding computationally demanding and hypothesis-driven^20^ choices during image processing - and model complex non-linear relations between voxels. One such model is the Simple Fully Convolutional Network (SFCN), a novel deep Convolutional Neural Network (CNN) that won the Predictive Analysis Challenge for brain age prediction in 2019 (PAC2019)^21,22^. While such deep learning approaches allow for the prediction of brain age with unprecedented accuracy, and can potentially help us identify idiosyncratic regional patterns of neurodevelopment and ageing at the individual level^23,24^, their complexity also comes with a risk of overfitting, namely finding patterns in the training data which do not generalize well to new, previously unseen, participants^25^.

Transfer learning, a deep learning technique widely used in other areas of applied machine learning research, has recently gained momentum in neuroimaging^26^. Here, learned intermediate representations can be shared between tasks, allowing a model to be transferred to a problem or a dataset it was initially not trained for^27^. This approach has arguably been one of the core developments underlying the practical success of deep learning in a range of computer vision problems^28^ by using models trained on general purpose datasets, typically ImageNet^29^, and fine-tuning them towards a wide array of tasks. Recent studies have shown that transfer learning yields promising results also for brain age predictions^30^ and clinical classifications based on MRI data^31,32^. This exemplifies the need for robust, pretrained models on massive multisite datasets, that can be translated to smaller clinical samples, and, ultimately, to individual cases in a clinical setting. Modelling brain age as a pre-training step has obvious advantages as age is a variable which is available in most current MRI datasets. Additionally, the representations learned by the brain age models, representing partly independent dimensions of age-related variance, could be of direct importance in individual level brain phenotyping.

In the present study we trained deep neural network models for brain age prediction on structural MRI data from 53542 healthy individuals between 3 and 95 years of age to test their ability to generalize, and demonstrate the downstream applicability and biological relevance of a properly generalizing model. We used the Simple Fully Convolutional Network with a softmax output (SFCN-sm) which predicts age as a discrete probability distribution, and compared its accuracy and generalizability with two proposed variants of the architecture; a regression variant (SFCN-reg), directly predicting continuous age, and a ranking variant (SFCN-rank) encoding age as an ordinal vector. Brain age delta was computed as the difference between the predicted and chronological age. To estimate the sensitivity and clinical relevance of our best model, we tested for associations between brain age delta and a range of clinical and biological phenotypes, and sociodemographic and lifestyle variables, in unseen data from a population sample. To further demonstrate the applicability of the model, we employed the pretrained SFCN-reg in a transfer learning context to predict case-control status for AD, MS, mild cognitive impairment (MCI), SCZ, mood disorders and psychotic disorders on datasets obtained from a range of MRI scanners. To promote transparency and reproducibility we have implemented an easy-to-use Keras interface for all the trained models, both the brain age and clinical predictors, and a pipeline for preprocessing images, available on our GitHub http://www.github.com/estenhl/pyment-public.

## Results

We compiled 21 publicly available datasets with T1-weighted MRI scans into a large and diverse imaging dataset (total N=53542; female N=27715; age range=3-95), and trained a Simple Fully Convolutional 3-dimensional CNN with 6 convolutional blocks and a softmax output layer (SFCN-sm, Figure 1b), as introduced in Peng et al^21^. We then proposed two alternatives for the prediction layer of this architecture, the first based on regression (SFCN-reg) and the second based on ranking^33^ (SFCN-rank) (see Methods for details). Due to the time-consuming process of model evaluation, we restricted our search to these three variants of the given architecture, and trained a handful of versions of each variant with different hyperparameter settings. To evaluate the generalization performance of the models, we divided our data at two levels; A reference dataset (Figure 1 and Supplementary Table 1) and an external dataset (Figure 2a and Supplementary Table 2). In the reference dataset, we evaluated the performance of the trained models on an independent test split from known scanners with an age distribution resembling the training split. We then tested model performance on the external dataset compiled from different sources, originating from scanners unseen by the models during training with a divergent age distribution.

**Figure 1:**
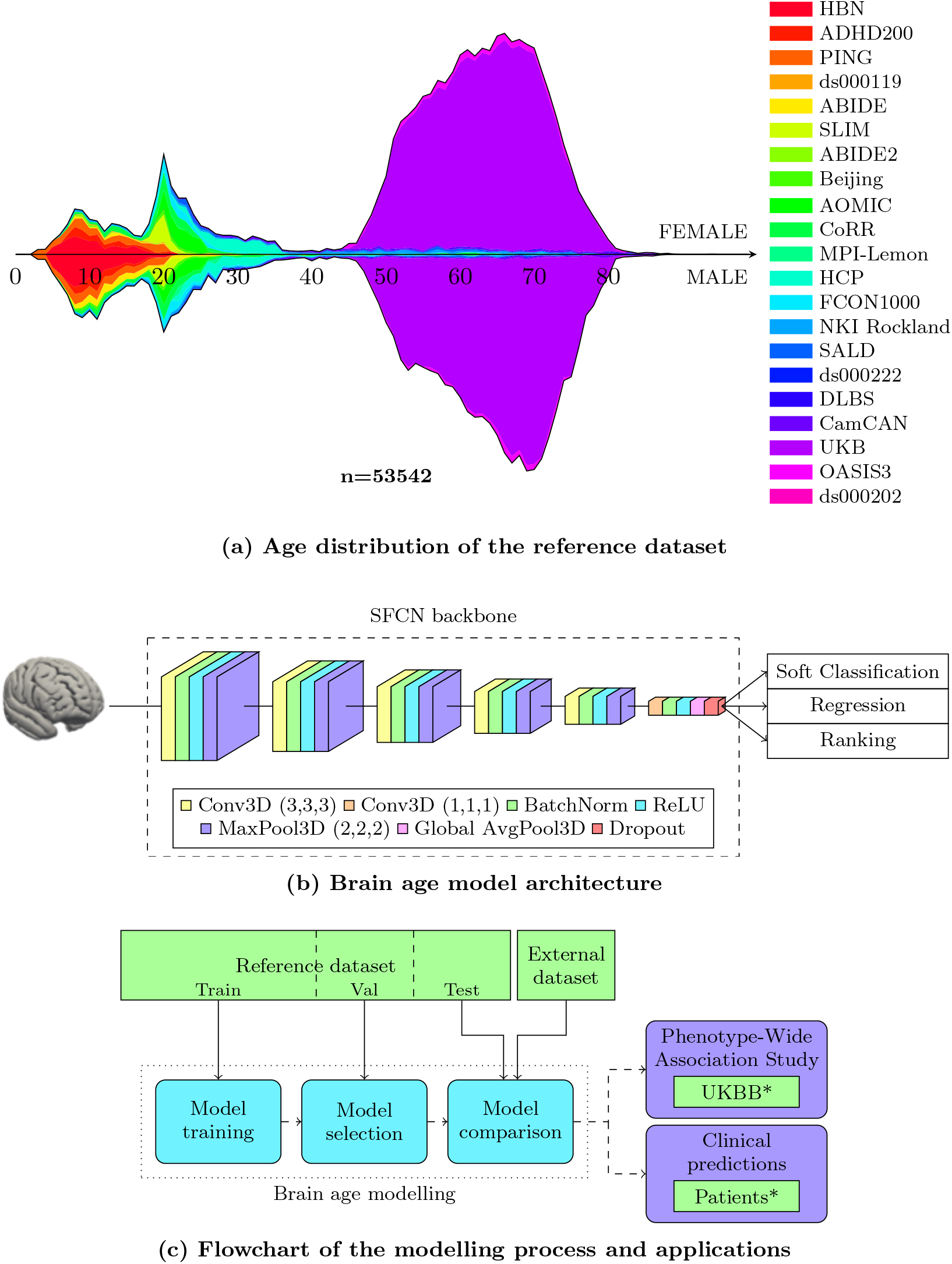
An overview of the dataset and models used for brain age modelling. (a) The reference dataset contains 53542 healthy participants from 21 datasets, with ages ranging from 3 to 95 years. For both sexes the age distribution is approximately bimodal, with the first large group of participants from 5 to 35 years originating from multiple studies, and the second from 45 to 80 mostly consisting of participants from the UKBB. (b) We implemented three model variants for predicting brain age, all based on the contest-winning SFCN architecture^21^. The architectures of the three variants differ only in the final prediction layer. All the models take minimally preprocessed T1-weighted MRI images as input. (c) The modelling process consisted of three steps, each utilizing a different portion of the reference dataset. In the final comparison we also employed an external dataset, coming from previously unseen scanners, to assess the generalization capabilities of the different models. Having found the best brain age model, we subsequently applied it in two applications: A phenome-wide association study (PheWAS), and a case-control comparison including several clinical conditions.

**Figure 2:**
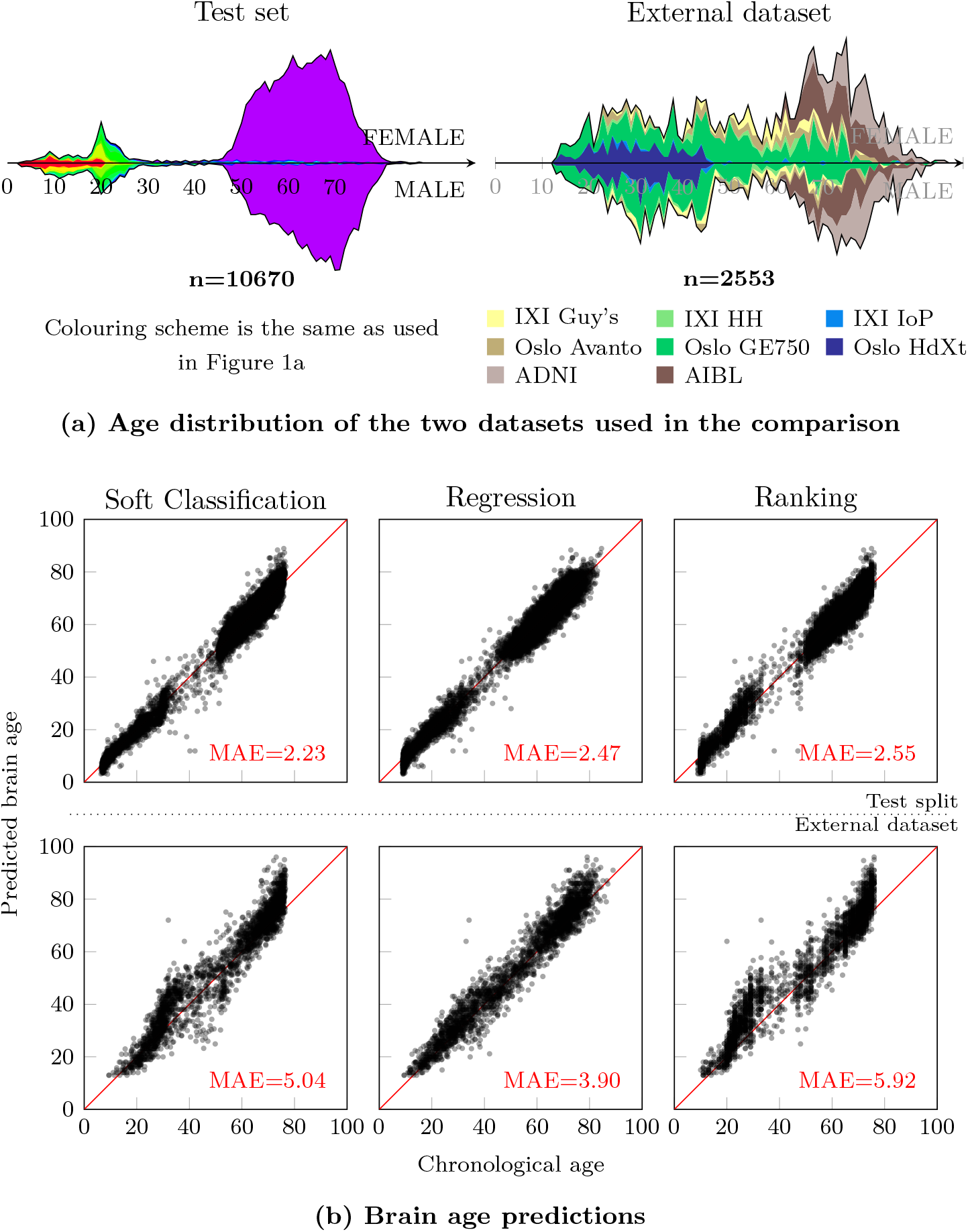
The two datasets used in the model comparison, and the predictive performance of the three model variants. (a) For comparing the models we employed two distinct datasets: A test set sampled from the reference dataset, and an external dataset. The former was drawn using stratification, and as a consequence has an age distribution closely resembling the one seen in the full reference dataset. The latter was compiled from a different subset of datasets, and thus was acquired from different scanners. The age range of the external dataset is somewhat narrower, spanning a region of 13 to 95 years, and has a more uniform distribution. (b) The six scatter plots display the predictions of a given model on the x-axis, against the ground truth age of the participants on the y-axis. The top row contains the predictions of the three model variants on the test set, and the bottom row the external dataset.

### Superior generalization performance of SFCN-reg

We used the training and validation sets to optimize and tune the models, and a conjunction of the test set and the external dataset in a final model comparison. For each model variant we trained three versions, with different hyperparameter settings, on the training data, and selected the best model based on the mean absolute error (MAE) on the validation set (Figure 1c and Supplementary table 8). We then compared the performances of the best version for each model variant on the test set. In this comparison the results mirrored those of the validation set, with SCFN-sm achieving the best result with an MAE of 2.23 years, followed by SCFN-reg with 2.47 years and SCFN-rank with 2.55 years (Figure 2b). Given the added complexity of including multiple datasets from a large range of scanners, we consider these results to be approximately on par with the MAE of 2.14 reported in the original SFCN paper^21^, and thus among the best performing models in the field^34^. Additionally, the heterogeneous origins of the dataset facilitate cross-site generalization, an essential property when training large multisite models^35^.

As a conclusive test of model generalization, we performed the same comparison on the external dataset, containing unseen data from different MRI scanners with an age distribution diverging from that of the reference dataset. SCFN-reg substantially outperformed the two alternatives, with an MAE of 3.90 compared to 5.04 and 5.82 for SFCN-sm and SFCN-rank respectively (Figure 2). While the performance of all models were lower on the external dataset, the extent of the generalization error was considerably different. When compared with MAEs from the test set, the average error of SFCN-reg increased by approximately half, while it more than doubled in SFCN-sm and SFCN-rank (Supplementary Table 9). This difference coincides with the architectural differences between the models: Where both the SFCN-sm and SFCN-rank used age-bins, with an output node for each age in its prediction range, SFCN-reg had a single output predicting a single continuous number. Therefore, the predictions of the SFCN-reg reflect a simpler combination of the learned representations in the preceding layer of the model, which we hypothesize may be the reason for the improved generalization performance. In the subsequent applications we use the SFCN-reg version that achieved the best MAE on the external dataset. Additionally, to facilitate cross-study comparisons, we have compiled a range of performance metrics for our three models on the external dataset in Table 1.

**Table 1:** Predictive performance of the soft classifiation model (SFCN-sm), the regression model (SFCN-reg) and the ranking model (SFCN-rank) on the external dataset, originating from scanners which has not been seen by the models during training. Mean Absolute Error (MAE), Root Mean Squared Error (RMSE), R and R2 are sensitive to the age range of the dataset, while normalized RMSE (nRMSE) and Relative Absolute Error (RAE) are not, facilitating comparisons across datasets.

| Model | MAE | RMSE | R | R<sup>2</sup> | mRMSE | RAE |
| --- | --- | --- | --- | --- | --- | --- |
| SFCN-sm | 5.04 | 6.51 | 0.961 | 0.903 | 0.078 | 0.26 |
| SFCN-reg | **3.90** | **5.11** | **0.975** | **0.940** | **0.061** | **0.20** |
| SFCN-rank | 5.92 | 7.54 | 0.959 | 0.870 | 0.090 | 0.31 |

Observing that all the models performed worse in the external dataset than in the test set, we performed post-hoc analyses to further understand the causes underlying the generalization problems. Specifically, we tried isolating two sources of generalization error: Differences in population, represented here by age and sex distributions, and differences in scanners and acquisition protocols. We approached this by resampling two artificial datasets, both with participants previously unseen by the models. First, we sampled a dataset with an “Unknown population”, with participants from the test set following the empirical age and sex distribution of the external dataset. Secondly, we created a dataset with “Unknown scanners”, sampling participants from the external dataset while following the distributions of the test set (see Methods for further details). Due to the stratification used in the initial train/validation/test split the latter set also directly matches the distributions of the training set. For each of these two new datasets we computed an MAE per model, which naturally fell between the MAEs on the test set and on the external dataset. While this approach is exploratory and inherently limited to the characteristics and actual data points making up our datasets, the results clearly indicate that the main driver of generalization error is the unknown scanners (Table 2 and Supplementary Figure 1) which had higher errors (MAE_US_) than the unknown population (MAE_UP_) for all the models. Additionally, these two sources of generalization error seem to work additively, with their sum closely matching the full generalization error observed in the external dataset. A final observation was that SFCN-reg handled both sources of error best, which is further evidence of its superior ability to generalize.

**Table 2:**
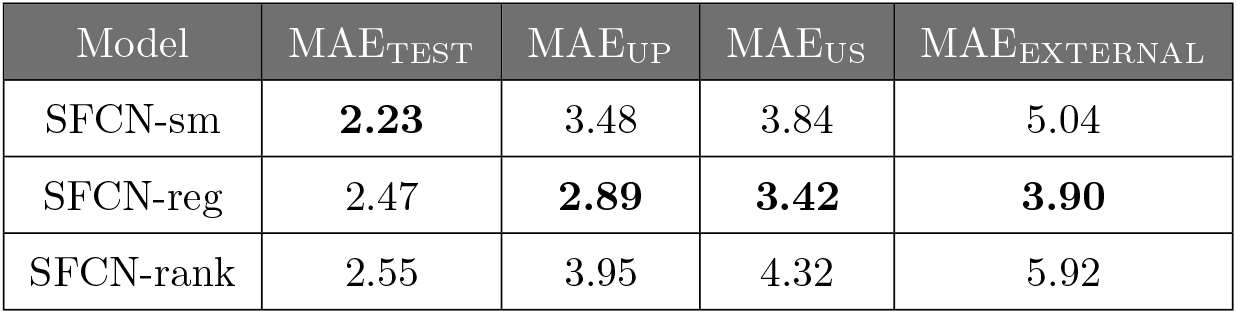
Mean Absolute Errors measured for the three model variants on the test set, drawn from the same distributions of scanners and ages as the training dataset, the “Unknown population” dataset (MAE_UP_) and the “Unknown scanners” dataset (MAE_US_), both representing a single source of generalization error, and the external dataset, different from the training set in both regards.

| Model | MAE<sub>TEST</sub> | MAE<sub>UP</sub> | MAE<sub>US</sub> | MAE<sub>EXTERNAL</sub> |
| --- | --- | --- | --- | --- |
| SFCN-sm | **2.23** | 3.48 | 3.84 | 5.04 |
| SFCN-reg | 2.47 | **2.89** | **3.42** | **3.90** |
| SFCN-rank | 2.55 | 3.95 | 4.32 | 5.92 |

### Brain age predictions associate with biological phenotypes and lifestyle factors

Next, we validated the relevance of the model predictions by correlating their deviations from chronological age with an array of phenotypes in a phenome-wide association study. We performed this analysis in the subset of the UKBB data that was not used for brain age modelling or validation (n=8066), and tested associations with all the biological phenotypes and lifestyle variables accessible, manually divided into thirteen thematic categories for interpretability (Supplementary Table 7). For each phenotype we computed a univariate correlation while correcting for age and sex^36,37^, and assessed its significance using a Bonferroni-corrected p-value threshold of *p <* 1.26 × 10^−4^ (see Methods). All continuous variables were standardized, such that their effect sizes denote the impact a one standard deviation increase has on the brain age delta. In general, our results corroborated several findings derived from previous studies using smaller samples (Figure 3a). We observed significantly higher delta in participants with high blood pressure (*β* = 0.41, *p* = 1.86 × 10^−7^), those currently on blood pressure medication (*β* = 0.54, *p* = 1.17 × 10^−10^), and a positive correlation with blood pressure readings (diastolic (DBP): *β* = 0.15, *p* = 2.53×10^−6^, systolic (SBP): *β* = 0.16, *p* = 3.30×10^−6^). The associations with the largest effects indicated higher delta in patients with a diabetes diagnosis (*β* = 0.74, *p* = 2.25 × 10^−7^) or diabetes-related eye problems (*β* = 1.78, *p* = 4.59 × 10^−7^). Among the biochemical measurements, significant associations with brain age delta were found for blood glucose levels (*β* = 0.23, *p* = 3.18 × 10^−11^), Insulin-Like Growth Factor-1 levels (*β* = −0.22, *p* = 5.81 × 10^−11^), glycated haemoglobin levels (*β* = 0.16, *p* = 7.97 × 10^−7^) and mean corpuscular volume (*β* = 0.13, *p* = 3.33×10^−5^). Associations with variables we categorized as related to diet and lifestyle were dominated by previous smoking, with a positive correlation with number of cigarettes per day (absolute pack years: *β* = 0.24, *p* = 8.55 × 10^−5^, pack years as proportion of age: *β* = 0.26, *p* = 2.41 × 10^−5^) and age stopped smoking (*β* = 0.25, *p* = 7.59 × 10^−5^). We also observed significant associations with average weekly beer and cider intake (*β* = 0.21, *p* = 1.13×10^−7^) and alcohol intake frequency (*β* = 0.12, *p* = 1.09 × 10^−4^), cereal intake (*β* = −0.16, *p* = 7.61 × 10^−7^) and participation in “Other group activity” (e.g. social activities not related to a sports or social club, religious group or adult education, *β* = −0.27, *p* = 5.91 × 10^−5^). Further, we observed a significant correlation with the number of people living in the participants household (*β* = −0.13, *p* = 8.20 × 10^−5^) and higher deltas in those born outside the United Kingdom and the Republic of Ireland (compared to the baseline group born in England, *β* = 0.62, *p* = 4.62 × 10^−6^). An overview of all the 394 associations can be found in Supplementary Table 12.

**Figure 3:**
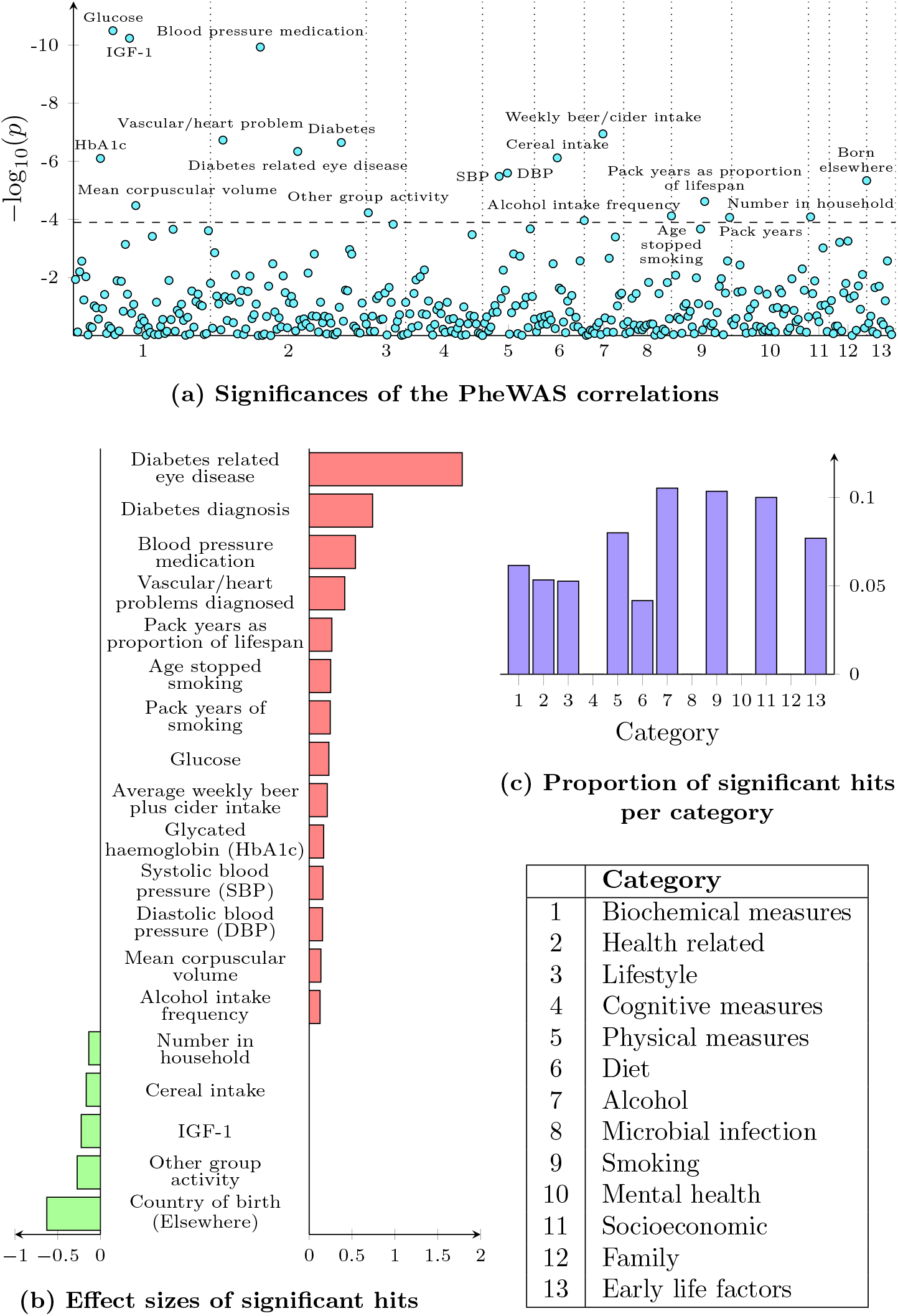
Associations between the brain age delta and a wide range of phenotypes. We correlated the brain age delta originating from the SFCN-reg with 394 phenotypic variables in the test set portion of the UKBB, categorized into thirteen categories. (a) A Manhattan plot visualizing the significances of the 394 associations. Using a Bonferroni-corrected threshold, 19 of these were found to be significantly associated with the delta. (b) The effect sizes of the significant associations. For binary variables the effect size expresses the mean difference between the groups, while for continuous variables it denotes the change in brain age delta associated with a one standard deviation increase. (c) For each of the thirteen categories we calculated the proportion of significant hits by dividing the number of significant hits within that category with the total number of variables in the same category.

### Transferring brain age predictions to developmental and degenerative brain disorders

For six different disorders we compiled a patient cohort and a matched control group, and calculated a brain age delta per participant based on the prediction from SFCN-reg (Supplementary Figure 4). In all control groups, the brain age prediction accuracy was approximately the same as for the full test set (MAEs=2.91-4.05, Supplementary Figure 3). Patients with MS showed significantly higher brain age estimates than their matched healthy controls (brain age group mean difference Δ = 4.42 years, *p* = 1.71 × 10^−22^, Cohen’s *d* = 0.87). A similar pattern was also observed for patients with AD (Δ = 2.81, *p* = 4.20 × 10^−20^, *d* = 0.59), MCI (Δ = 2.13, *p* = 1.25 × 10^−15^, *d* = 0.46) and SCZ (Δ = 1.40, *p* = 4.29 × 10^−5^, *d* = 0.34). For the individuals with mood disorders (MOOD, see Methods) this difference was the smallest (Δ = 0.64, *p* = 0.04, *d* = 0.17), while the difference was not significant for patients with a mix of psychotic diagnoses (PSY) (Δ = 0.74, *p* = 0.15, *d* = 0.20). Both the relative ordering of the disorders in terms of group difference, the magnitude of the disparities, and the observed significance resemble a previous study^8^ using a different model based on a smaller dataset.

**Figure 4:**
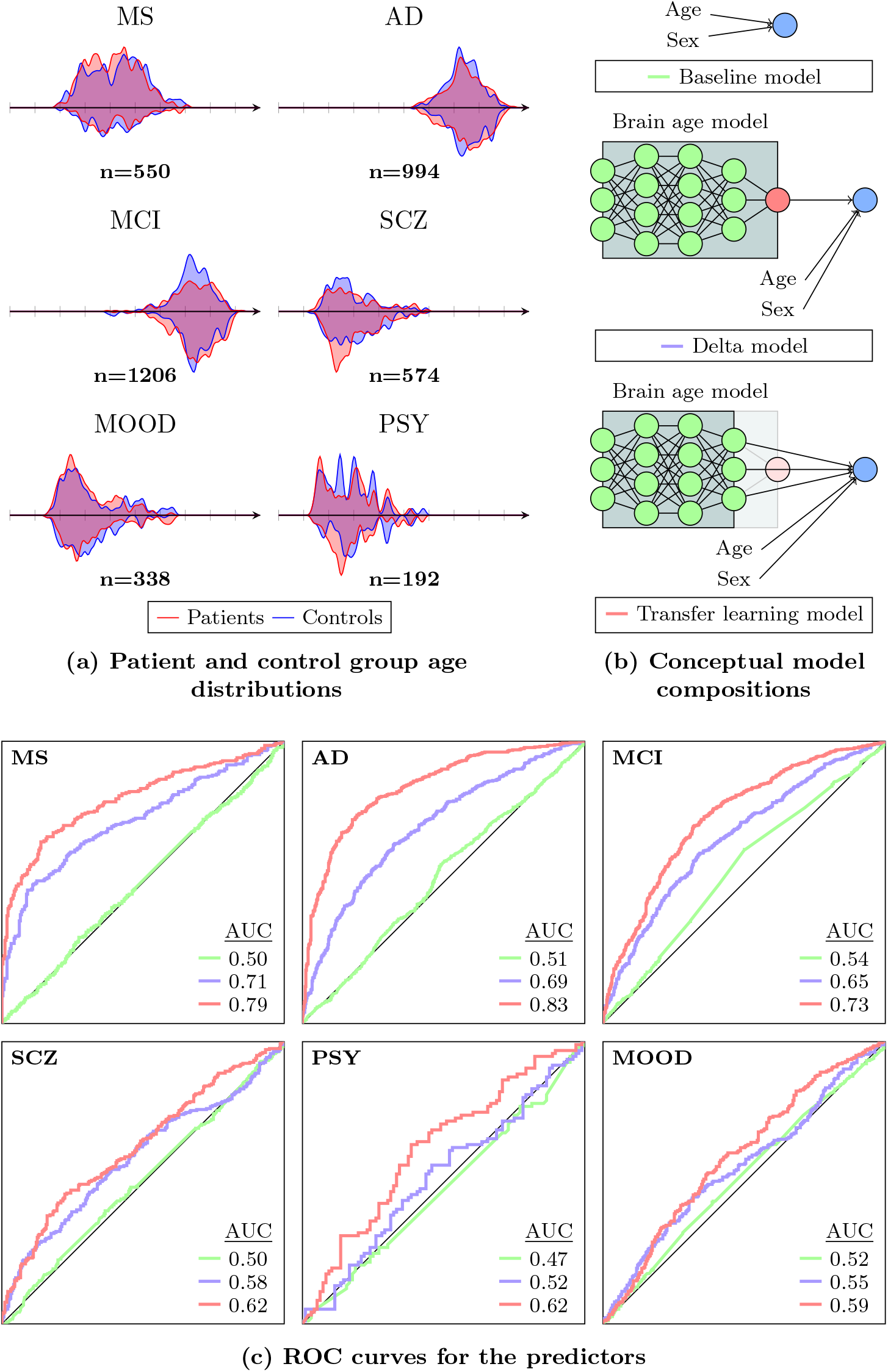
The datasets and models used for clinical predictions, and an overview over their performance. We applied the best performing SFCN-reg in a clinical setting by training binary classifiers to discriminate between patients and controls for multiple common brain disorders, using different levels of information from the brain age model. (a) To avoid biases we used a strict matching procedure, drawing a set of controls for each scanner-specific patient dataset matching its empirical age and sex distribution. (b) We trained three logistic regression models for each disorder to compare their performance. The first used age and sex as predictors in a baseline model, the second included the brain age delta, quantifying its value as a clinical predictor, and the third used internal features of the brain age model in a transfer learning setting. (c) For each disorder we compared the binary models using AUCs, indicating the information content of the predictors in relation to the given disorder.

To demonstrate the predictive power of our best performing pretrained brain age model for clinical conditions, we trained multiple binary classifiers to predict whether a participant had a diagnosis or not (Figure 1c, Figure 4 and Methods). We used logistic regression models with an *l*_1_-penalization for this purpose (LASSO models), optimized via a nested cross validation procedure (Supplementary Figure 5), and started with a baseline model classifying participants based only on age and sex. The second model included the brain age delta originating from the brain age prediction of SFCN-reg, and the third model replaced the brain age delta by 64 features encoded in the second to last layer of the same model (Figure 4b and Methods). Across the six disorders, the baseline models achieved an area under the receiver operating curve (AUCs) ranging from 0.47 to 0.54, indicating that our matching procedure was satisfactory. Using the second set of models, quantifying the predictive power of the brain age deltas in Supplementary Figure 4, greatly improved the prediction performance when compared to the corresponding baseline models for MS (AUC=0.71 vs. 0.50), AD (0.69 vs. 0.51) and MCI (0.65 vs. 0.54); but to a minimal extent for SCZ (0.58 vs. 0.50), MOOD (0.55 vs. 0.52), and PSY (0.52 vs. 0.47). A third set of classifiers were implemented in a strict transfer learning context, utilizing the 64 features of the second-to-last layer of SFCN-reg as predictors (Methods). These features represent high-level, data-driven abstractions of the brain imaging data, and underlie the singular brain age prediction. We refer to this variant of transfer learning as strict because we kept the weights of the initial brain age model locked while optimizing for the new binary objective, which in turn allow us to keep treating these as ageing features and thus promote interpretability. While this complicates contextualizing the performance of our models in terms of existing case-control classifiers, it gives us an indication of the information content of these learned features. This third set of models improved AD prediction substantially (AUC=0.83), and also were notably better for MS (0.79), MCI (0.73) and PSY (0.62), while only a marginal improvement was observed in SCZ (0.62) and MOOD (0.59). Overall, our results show that our brain age model can be transferred to make case-control predictions of these common clinical brain disorders.

## Discussion

Brain maturation and ageing, and its interactions with clinical brain disorders and conditions, are complex processes with pivotal environmental and genetic contributions^38,39^. Brain age prediction and the accompanying brain age delta has the potential to provide intuitive and useful measures for summarizing individual brain aberrations. However, technical differences between studies, e.g. the use of different scanners and MRI scan parameters, have represented challenges for the direct generalization and applicability of brain age models based on large training sets. We assembled a large and diverse neuroimaging dataset to train multiple state-of-the-art deep learning models for brain age prediction, and extensively tested their ability to generalize, and their sensitivity to various common brain disorders. Our best model, the SFCN-reg, showed superior performance on an external dataset with differing scanners and age distributions compared to that of the training set, enabling it for applications in other datasets. We then demonstrated the relevance of the model predictions by showing associations between the brain age delta and a range of complex human traits and health outcomes in a population sample. Lastly, we transferred the trained SFCN-reg to clinical data in a transfer learning setting, showing that both the brain age delta and the internal features learned by the model have predictive value when differentiating between controls and patients with common brain disorders.

In our experiment the SFCN-reg outperformed the other models in terms of generalization performance. The age prediction accuracy on the test data (MAE=2.47) is among the best in the field^34^. Crucially, the prediction accuracy on data coming from unknown scanners (MAE=3.90) fares very favourably when compared to other studies attempting to transfer between datasets of similar size and complexity^30,32,34,40,41^. Using data from different scanners, protocols and populations in neuroimaging comes with the problem of modelling the effects of these appropriately^18,42^. Previous brain age studies have often explicitly included a scanner term in the modelling or corrected the computed brain age for various biases^43^. Recent approaches have tried to address this problem directly via specific deep learning paradigms^44^. Our results show that given sufficiently large and heterogeneous training data, deep CNNs achieve state-of-the-art performance for brain age predictions even when scanner effects are not explicitly modelled, and more importantly that this performance translates to scanners and protocols that are unknown to the model. This suggests that the representations learned by the model are dominated by age-related variance, not scanner-dependent artefacts, an extension of the model robustness shown in earlier studies^19^. From a practical point of view this suggests that our trained model may be employed in other applications in new datasets, without the need for retraining or applying corrective procedures.

Evaluating the biological relevance of the brain age predictions is essential to further understand and trust these models. Therefore, we correlated brain age delta obtained from the SFCN-reg with a wide range of phenotypes in a subset of the UKBB not used for model training. When applying a Bonferroni-corrected threshold, we found nineteen significant hits spread across seven of our thirteen categories. Almost all of these have in earlier studies been found to have a relationship with ageing, either generally or specifically in the brain, or with brain health and/or cognitive function: Glucose level^45^, Insulin-like Growth factor-1^46^, and glycated haemoglobin^47^ are known to change with age, and corpuscular volume has been associated with cognitive functioning^48^. Lifestyle factors involving alcohol and smoking impact various biological and bodily ages^45^, including that of the brain^49–51^. Elevated blood pressure and other cardiovascular risk factors have established associations with increased brain age^50,52^, and an increase in predicted age has been observed in patients with diabetes^53^. In sum, this analysis shows that our model makes biologically meaningful predictions. Further, transferring the model to unseen datasets comprised of patients with different clinical conditions and matched healthy peers revealed both high accuracy in terms of age prediction and higher deltas among patients with brain disorders, in line with previous studies^8^. Importantly, the data in the case-control datasets were obtained from scanners not included in the training set, supporting that the model generalizes to previously unseen scanners; a highly valuable asset.

Compared to the all-in-one brain age delta, a single number describing the difference between apparent and chronological age, our results showed increased predictive value for MS, AD and MCI when using the internal representations of the SFCN-reg underlying the brain age predictions. This supports the view that for some applications the constituent components of the singular brain age delta are relevant beyond the age prediction alone^54^. The innate ability of deep neural networks to form abstractions of the brain at different spatial resolutions throughout the layers of the model may help disentangle individual differences in neurodevelopmental and age-related processes related to complex disorders and traits. Furthermore, we see this result as evidence that deep learning models trained to predict age in large multisite datasets constitute excellent starting points for transfer learning, which can subsequently be fine-tuned to a variety of tasks.

There is an ongoing discussion in the field on whether brain age models that are precise, or those that allow for sufficient variance in their single-subject predictions, are the most useful in a downstream analysis of behavioural and clinical traits^32,55,56^. An argument for a model which allows for more variation (a looser fit) is that this would more accurately depict brain age as a complex process which appears differently in different individuals. One challenge with this approach is that the brain age delta is a residual, and recognizing what portion of this error comes from biological variation and what is modelling imprecision is practically unfeasible. As more complex models such as deep CNNs become competitive for brain age modelling, it becomes possible to minimize the overall model error, including the methodological portion, while still allowing the model to accurately represent the necessary biological variability internally.

There are some limitations of the present study which we acknowledge here. Given the computational cost of training complex deep learning models on such a large dataset we restrained our study to a limited number of possible models, both in terms of model architectures and hyperparameter settings. With further increases in sample sizes and diversity, deeper architectures may be sensible, as the risk of overfitting is directly alleviated by the larger datasets. While we refer to our dataset as diverse, most participants included in the study are of white European background. However, this limitation is not specific to the present study, and global collaborations are needed to build models that generalize across cultural and genetic backgrounds^57^. Relatedly, the current investigation included T1-weighted MRI data only. Future integration of information spanning various imaging modalities may increase both age prediction accuracy and the sensitivity and specificity of various biological and clinical traits and conditions^11,52,54,58^. To evaluate the clinical relevance of the brain age model in the context of disorders we employed transfer learning in a strict way, by training LASSO models on top of the SFCN architecture. This setup increases the interpretability of the transfer process as the lower-level representations of our model used as predictors capture age-related features of the brain. In turn, this allows for the interpretation that illnesses that are predictable by the model must also rely on these representations, and thus implicitly relate to age. There are multiple steps which could have been taken instead to maximize predictability, a natural starting point is to fine-tune the entire model^32^. Lastly, in order to reach broad adoption of these models and, ultimately, approach clinical usability, a better understanding of the regional patterns driving the prediction, their specific biological significance and how it changes across time and contexts is needed^59^.

In conclusion, we have trained multiple variants of a deep neural network to predict brain age on a large and heterogeneous sample of raw structural MRI data, and observed distinct differences in their ability to generalize to unseen samples and scanners. The predictions of our best model were linked to biochemical biomarkers, cardiovascular risk factors, smoking and alcohol intake, among others. Using transfer learning, we demonstrated that clinical conditions with a neurodevelopmental or neurodegenerative aetiology were predictable by our model, initially trained to predict age. Jointly, these findings add to the growing literature documenting the tremendous potential of advanced techniques for statistical learning to decode biologically and clinically relevant information from brain MRI scans.

## Methods

### Data

All data sets used in the present study have been obtained from previously published studies which have been approved by their respective institutional review board or relevant research ethics committee.

### Reference dataset

The reference dataset used for training the brain age models was T1-weighted MRI scans derived from 21 non-overlapping and publicly available datasets (total n=53542; female n=27715) of healthy individuals, with ages ranging from 3 to 95 years (Figure 1 and Supplementary Table 1). The age distribution can be seen in Figure 1a. The younger age-range (3-30 years) was mainly composed of participants from multiple different datasets. Though the older age-range (40-80) also included multiple datasets, UKBB accounted for most of these participants. The most sparsely populated age-ranges were in the very young (147 participants with age ≤ 5), very old (17 participants with age ≥ 85), and in midlife (42 participants with 35 ≤ age *<* 45). For each of the datasets, participants that had one or more psychiatric, neurological and/or other relevant diagnoses (Supplementary Table 4), and those withdrawn from the respective study were excluded before model training. In addition, for participants having multiple brain scans, the baseline data were used, such that in the final dataset each data point represents a unique participant drawn from a normative population.

### External dataset

To evaluate the generalizability of our trained brain age prediction model, an external dataset was collected (Supplementary Table 2). This dataset included the IXI project (Supplementary Table 5) and healthy controls from the datasets underlying the clinical data described below (total n=2553). Importantly, the external dataset contained images generated by scanners not used in the reference dataset, and subsequently unknown to the models during training and validation. This dataset can be seen as having a more uniform age distribution (Figure 2a), meaning that our test would capture whether any given model relies too heavily on information observed in the training data.

### Clinical data

The clinical data consisted of six patient cohorts diagnosed with MCI, AD, MS, SCZ, a mix of psychotic diagnoses and mood disorders, where the latter was a combination of two cohorts with depression and bipolar disorder (Figure 2a and Supplementary Table 3). The individual cohorts were compiled from ADNI, AIBL, and multiple scanners at the Oslo University Hospital (Supplementary Table 5 and 6). In addition to the patients, we used healthy controls from the same scanners in the external dataset to create matched control groups for the clinical predictions.

### Quality control

To ensure data quality, we executed a quality control (QC) pipeline, consisting of checking whether any of the image preprocessing steps failed, and a manual control via visual inspection. To take advantage of as much data as possible this manual control was lenient, removing samples where either a significant portion of the brain was missing, or where the orientation of the head was dramatically off, and resulted in dropping only 39/53581 participants in the reference dataset (0.07%) and 1/2554 participants in the external dataset (0.03%).

### Image preprocessing

We first performed skull-stripping with the FreeSurfer 5.3 auto-recon pipeline^60^ to produce a brain-mask, minimizing the amount of non-brain information in the data, then reoriented the images to the standard FSL^61^ orientation using fslreorient2std. The resulting images were linearly registered to the MNI152 space using FLIRT^62^ with linear interpolation and the default 1 mm FSL template (version 6.0). We cropped away borders of [6:173, 2:214, 0:160] voxels, in the sagittal, coronal and axial dimensions respectively. This cropping yielded the smallest cuboid with marginal loss of brain-related information across the dataset, minimizing the memory footprint of the models during training. As a last preprocessing step the voxel intensity values of all brain images were normalized to the range [0, 1].

### Brain age models

The state-of-the-art network architecture, the Simple Fully Convolutional Network (SFCN)^21^, was implemented as the backbone in all our brain age models. The SFCN architecture consists of a VGG^63^-like structure, with five repeated convolutional blocks, each with a three-dimensional convolutional layer with a filter size of (3, 3, 3), a batch normalization layer, rectified linear activation function (ReLU) activation, and a max-pooling layer with a pooling size of (2, 2, 2) (Figure 1b). The model then has a channel-wise convolutional layer, a last batch normalization layer and a global average pooling layer. From this backbone we defined three end-to-end variants for brain age prediction: The original soft classification model (SFCN-sm), a regression variant with a single output node with a linear activation (SFCN-reg) and a ranking model from the general age-regression literature (SFCN-rank)^33^, an approach which has also been successful for brain age predictions^64^. All the model definitions rely on a matrix *X* of dimensions [*N, h, w, d*] containing MRI images as input, an N-dimensional vector age=[age_0_, age_1_, …, age_*N* −1_] containing the ground truth ages of the participants to compute its loss, and are ultimately able to produce an N-dimensional vector aĝe = [aĝe_0_, aĝe_1_, …, aĝe_*N* −1_] with a single brain age prediction per participant (although this is not necessarily the direct output of the model).

### SFCN-sm

The soft classification variant formulates the age regression problem as a multiclass classification problem, by having *m* = ⌈max(age) ⌉ − ⌊min(age) ⌋ output neurons, where ⌈· ⌉ and ⌊ ·⌋ denote the ceiling and floor operators respectively. It is denoted as soft because it uses a target vector per participant generated by a normal distribution centered around the ground truth age, instead of the one-hot encoding used in regular classification

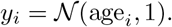

The predictions of the model are similarly a vector of length m with a softmax activation

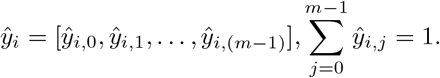

The loss for a single datapoint is the Kullback-Leibler divergence between the two vectors

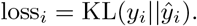

The final age-prediction of a participant is calculated as a weighted sum of the prediction vector

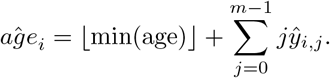

### SFCN-reg

The regression variant has a single output neuron predicting a single value *ŷ*_*i*_ per participant, limited to the range (min(age), max(age)) with a bounded ReLU activation. The value of *ŷ*_*i*_ can be used directly as the predicted age for a participant, *aĝe*_*i*_ = *ŷ*_*i*_. During training, the model optimizes the mean squared error.

### SFCN-rank

The ranking model formulates the age regression problem as a set of binary “*Is participant X older than age y?* “-questions. Like the soft classification model, the model has *m* = ⌈max(age)⌉ − ⌊min(age)⌋ output neurons, each representing one such binary question. The target vector for a participant is a binary vector of length *m*, with a 1 in bins corresponding to ages younger than the participants age and 0 in the rest.

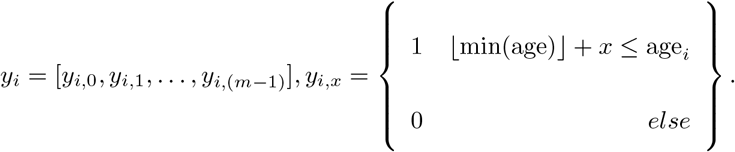

For each participant the model predicts a vector of length m, where each output neuron is limited to the range [0, 1] by a sigmoid activation

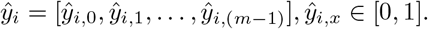

The model optimizes the mean binary cross entropy across all the output neurons

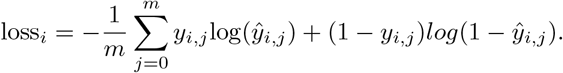

To calculate a predicted age for the model we sum up the number of age bins for which the model predicts that a participant is older than the given age (it is worth noting that this limits the model to integer predictions)

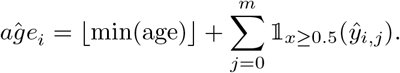

### Brain age model training and comparison

All brain age models were trained on 2 NVIDIA V100 GPUs with 32GB memory, using the Keras^65^ interface of Tensorflow 2.3^66^ on top of cuda 10.0. Using a batch size of 14 the models took approximately 1 second per step, translating into roughly 45 minutes per epoch or about 2.5 days per full training session. To train the brain age models, 80% (n=42829) of the reference dataset was used for model building (training and validation) and 20% (n=10713) for testing. Among the data for model building 80% (n=34285) and 20% (n=8544) were used for training and validation of the models, respectively (Figure 1c and Supplementary Table 1). Before these splits, the data was stratified by age and original study to ensure that all subsets had resembling age distributions and came from multiple scanners. Given the great computational cost of model training, determining optimal hyperparameter values by searching over the full configuration space for each model is impractical. Instead, we employed post-hoc heuristics, i.e., tweaking the models based on previous runs. For each training run we trained the model from scratch (with randomly initialized parameters) for 80 epochs, optimized by vanilla stochastic gradient descent, employing an annealing, step-wise, learning rate schedule. This schedule had three steps, reducing the learning rate by a factor of 3 after epochs 20, 40 and 60. The initial learning rate was found independently for each model variant using a learning rate sweep^67^ (Supplementary Figure 7). We used mean absolute error (MAE) on the validation split to determine the best epoch for each run. We also report RMSE, R, R^2^ for all models in Table 1, and to enable comparisons with other studies with possibly different age ranges the normalized measures normalized RMSE (nRMSE)

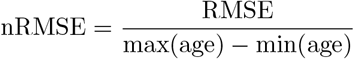

and Relative Absolute Error (RAE)

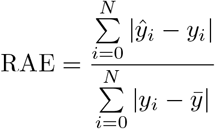

The first model we trained was SFCN-sm with the hyperparameters specified in the original SFCN paper^21^. Seeing that this model was underfitting we relaxed the regularization for a second run of the same model, and subsequently a third. The two hyperparameters we tuned in this process were the weight decay, and the dropout rate between the two final layers of the model. Having trained three soft classification models, we moved on to train three regression models and three ranking models using these same heuristics (Supplementary Table 8). To select a candidate model for each variant we compared the MAEs on the validation split. In the final model selection, we compared the MAEs of the candidate models for each variant on the test set and the external dataset.

### Post-hoc generalization analysis

To study the causes of the differences in generalization, we designed an experiment to isolate the underlying sources of this error. Based on previous knowledge of the problems of new scanners, and the predictions of the models at different ages (Figure 3) we specifically targeted two possible sources: Differences in population, represented by different distributions of age and sex, and data coming from unknown scanners. For each source we sampled an artificial, bootstrapped dataset based on our existing data. For the “Unknown population” dataset we sampled participants from the test set (originating from the reference dataset), to match the empirical age and sex distribution of the external dataset. Similarly, for the “Unknown scanners” dataset we sampled participants from the external dataset (coming from scanners unknown to the model) to match the age and sex distribution of the test set (and thus also the training set). The idea behind both datasets is to isolate a single source of generalization error. For robustness, we bootstrapped each of these two artificial datasets 100 times and reported the mean MAE achieved by the different models. Each sample was drawn probabilistically, with replacement, with the probability of drawing participant *x* of age *x*_*a*_ and sex *x*_*s*_ from dataset source based on the age and sex distribution of dataset target given by

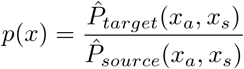

where 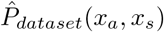 denotes the proportion of participants in dataset *dataset* with age *x*_*a*_ and sex *x*_*s*_. In “Unknown population”, the test set plays the role of source and the external dataset is the target, while this is switched for “Unknown scanners”.

### Phenome-wide association study

In the PheWAS we calculated the univariate correlation between the brain age delta and 402 phenotypic variables from the UKBB, manually divided into thirteen thematically defined categories for interpretability (Supplementary Table 7). We performed this analysis in the UKBB portion of the test split (n=8066), and used all the variables available to us at the time. We encoded all phenotypic variables according to the PHESANT^68^ datatypes, and removed non-informative levels (Supplementary Table 10) based on the UKBB coding schemes. Additionally, we re-coded the ordinal variables as categorical or continuous by hand (Supplementary Table 11). Variables which were impossible to model (i.e. singular or all missing values) were discarded. We then fitted a linear model per variable, modelling the delta as a function of the given covariate, age and sex, using the Python statsmodels API^69^. All continuous variables were standardized using a z-score normalization pre-modelling, such that the reported effect sizes refer to the change in brain age delta associated with a one standard deviation increase in the given variable. For assessing the significance of the associations, we computed a Bonferroni-corrected p-value threshold *p*_*bonferroni*_ = 0.05*/*394 = 1.12 × 10^−4^.

### Transfer learning to clinical samples

In the transfer learning analysis we trained multiple binary models to predict whether a participant had a given diagnosis or belonged to the control group, based on various levels of information from the brain age model. For this purpose, we used the clinical dataset which was previously unseen by the model, and matched controls groups drawn from the external dataset used in the generalization test.

### Control matching

To avoid biases in the case-control datasets we drew the subsets of controls independently for each scanner in the patient dataset, matching empirical distributions of age and sex in the corresponding case subset. For each disorder, for each scanner, the control group was created by drawing *n*_*case*_ controls, without replacement, using the sampling procedure described for the post-hoc generalization analysis.

### Feature extraction

To generate the feature vectors for each participant we used the trained SFCN-reg model as an encoder, up until and including the global average-pooling layer. Running a single MRI through the model up until this point results in a 64-dimensional vector representing the original image. Each of the dimensions *n* = [0, …, 63] in this space represent a high level feature of the brain, and each participant *X*_*i*_ = [*X*_*i*,0_, …, *X*_*i*,63_] is encoded as a point in this space. When used as a predictor in the subsequent modelling phase, each of these 64 dimensions were treated as an independent variable.

### Modelling

For each disorder we compared three different LASSO models, all trained and evaluated using the following general procedure, but on different covariate sets. We first stratified the given dataset on disorder, age and sex, respectively, and split it into 5 folds. We performed an outer cross-validation over these splits to allow us to have an out-of-sample prediction for each participant. When training a model on the training folds we performed an inner cross-validation to find the optimal value of the penalty parameter *λ*. The nested cross-validation procedure is illustrated in Supplementary Figure 5. Having found *λ*, we retrained the model on all the data from the four training folds. The models were implemented using sklearn’s LogisticRegression^70^ with an *l*_1_-penalty. Having the out-of-sample predictions for all the participants allowed us to calculate and compare AUCs based on the entire case-control dataset for the given disorder.

In addition to training the LASSO-model based on the brain age features, we trained an MLP using Keras with the same inputs. We did not optimize hyperparameters for this model, but observed similar results as the best LASSO models with the initial configuration (Supplementary Figure 6). The main benefit of the MLP is that it does not require a two-step process for the clinical prediction models, first processing the images with the encoder and then doing a prediction via a separate API, but can be implemented as an end-to-end binary classifier in Keras taking MRIs as inputs, and thus are more accessible for use by others.

## Data Availability

The raw data incorporated in this work were gathered from various resources. Material requests will need to be placed with individual principal investigators. A detailed overview of the independent datasets, and their origins, is provided in the supplementary information.

## Data availability

The raw data incorporated in this work were gathered from various resources. Material requests will need to be placed with individual principal investigators. A detailed overview of the independent datasets, and their origins, is provided in Supplementary Table 5.

## Code availability

All of the trained brain age models, the end-to-end clinical predictors, and a pipeline for preprocessing images is released in our GitHub repo at http://www.github.com/estenhl/pyment-public

## Acknowledgements

This work was funded by the UiO:LifeScience Convergence Environment (project: 4MENT), The Research Council of Norway (302854, 223273, 249795, 298646, 300767), the South-Eastern Norway Regional Health Authority (2014097, 2016083, 2018037, 2018076, 2019101), Stiftelsen Kristian Gerhard Jebsen, ERA-Net Cofund through the ERA PerMed project IMPLEMENT, and the European Research Council under the European Union’s Horizon 2020 research and Innovation program (ERC StG, Grant 802998) and the Wellcome Trust grant (215698/Z/19/Z). T.W. gratefully acknowledges support from the European Unions Horizon 2020 research and innovation programme under the Marie Sklodowska-Curie grant agreement No 895011. A.F.M. gratefully acknowledges support from the European Research Council (ERC, grant number 10100118) and the Dutch Organisation for Scientific Research (016.156.415). Part of the computation was performed on the Norwegian high-performance computation resources, sigma2 (NN9767K/NS9769K). Data used in preparation of this article were obtained from the Alzheimers Disease Neuroimaging Initiative (ADNI) and the Australian Imaging Biomarkers and Lifestyle Study of Ageing (AIBL) databases (adni.loni.usc.edu), and the Pediatric Imaging, Neurocognition and Genetics (PING) study database (chd.ucsd.edu/research/ping-study.html, now shared through the NIMH Data Archive (NDA)). The investigators within the ADNI, AIBL, and PING studies contributed to the design and implementation of ADNI, AIBL, and PING or provided data but did not participate in the analysis or writing of this report. This publication is solely the responsibility of the authors and does not necessarily represent the views of the National Institutes of Health or PING investigators.

## Notes

### Competing Interest Statement

The authors have declared no competing interest.

### Author Declarations

All data sets used in the present study have been obtained from previously published studies which have been approved by their respective institutional review board or relevant research ethics committee. A detailed overview of the data sources can be found in the supplementary information.

