## Supplementary Figures and Tables 1-9 for "Deep neural networks learn general and clinically relevant representations of the ageing brain"

Leonardsen et al.

### List of Supplementary Figures

1 Results of the model comparison on the resampled datasets representing a breakdown of the sources of generalization error. . . . . . . . . . . . . . . 2
2 Predictions of SFCN-reg on the external dataset, separated into the different scanning sites . . . . . . . . . . . . . . . . . . . . . . . . . . . . . . . . 3
3 Predictions of SFCN-reg on the control sets used in the clinical classifiers 4
4 Brain age delta distributions for the individual patient cohorts versus their individually matched control groups . . . . . . . . . . . . . . . . . . . . . 5
5 The nested cross-validation scheme used for the clinical predictors . . . . 6
6 A comparison of the AUCs of the LASSO and CNN clinical predictors . . 7
7 Results of the learning rate sweep for the three model variants . . . . . . 8

### List of Supplementary Tables

1 The reference dataset . . . . . . . . . . . . . . . . . . . . . . . . . . . . . 9
2 The external dataset used for generalization testing . . . . . . . . . . . . . 10
3 Patient cohorts used in the clinical classifiers . . . . . . . . . . . . . . . . . 11
4 Exclusion criteria for the participants in the reference dataset . . . . . . . 12
5 Sources of all the datasets used in the study . . . . . . . . . . . . . . . . . 15
6 Scanning protocols used in the datasets . . . . . . . . . . . . . . . . . . . 16
7 Columns used in the PheWAS . . . . . . . . . . . . . . . . . . . . . . . . . 19
8 The hyperparameter settings tested for each of the three model variants . 20
9 Results of the final model comparison . . . . . . . . . . . . . . . . . . . . 21

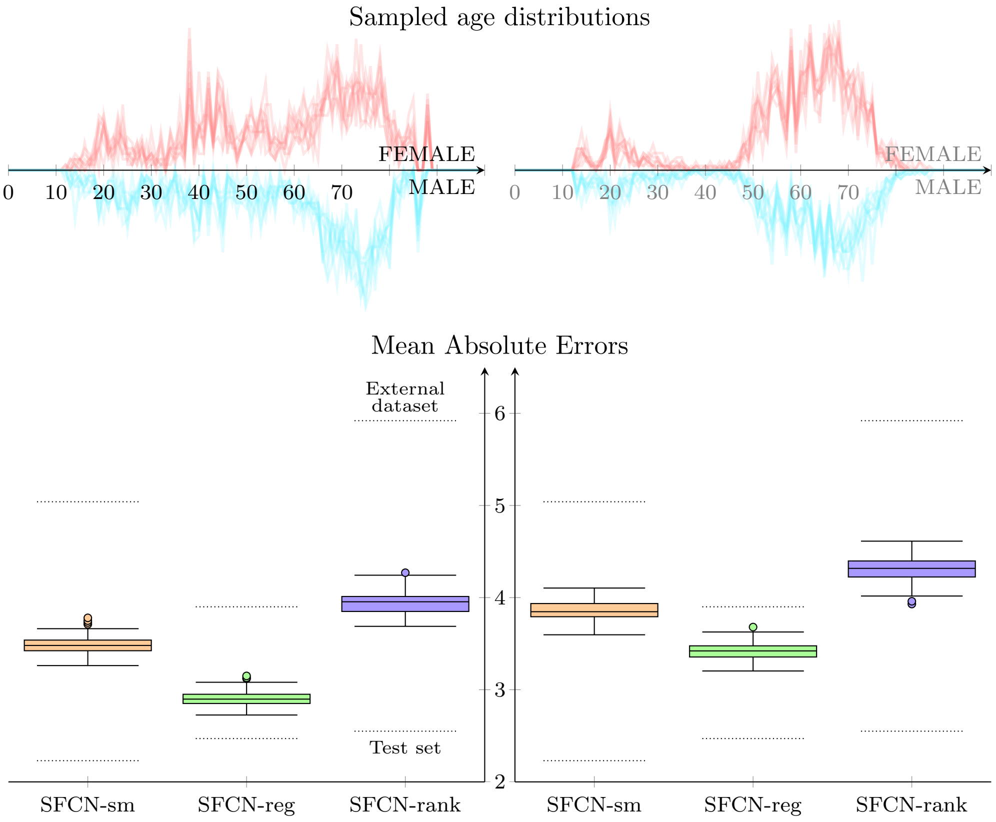


**(a) Unknown population** **(b) Unknown scanners**

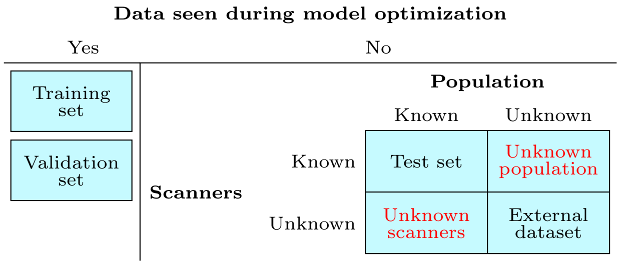


**(c) Sources of generalization error**

**Supplementary Figure 1: The results of the model comparison on the resampled datasets representing a breakdown of the sources of generalization error.** (a) For the ”Unknown population” dataset we drew 100 bootstrapped subsets from the test set matching the empirical age and sex distributions of the external dataset, and tested the performance of SFCN-reg. (b) For the ”Unknown scanners” dataset we similarily drew 100 bootstrapped subsets from the external dataset, matching the distributions of the test set.(c) A conceptualization of the assumed sources underlying the generalization error and the characteristics of the various datasets.

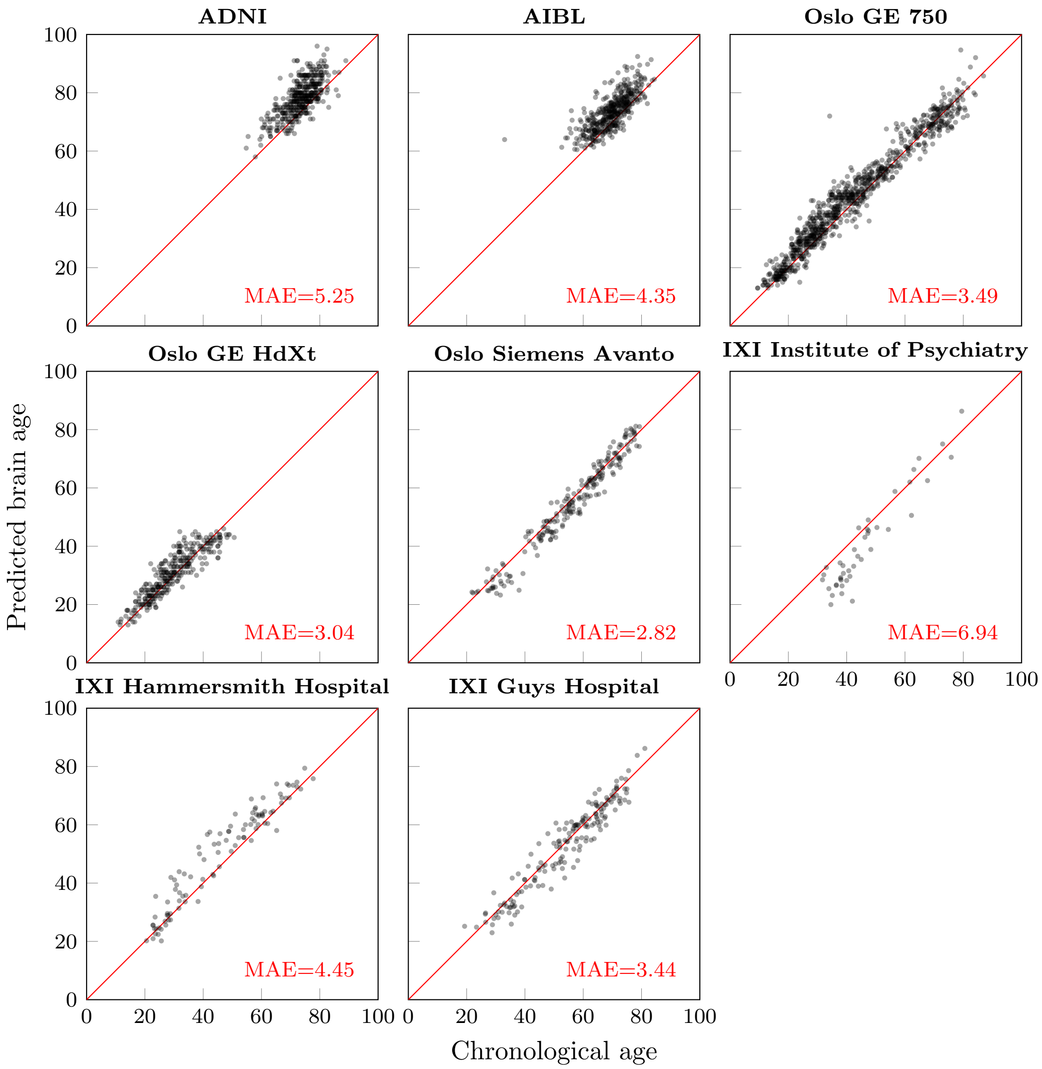


**Supplementary Figure 2: Predictions of SFCN-reg on the external dataset, separated into the different scanning sites.** Due to the large number of scanners, AIBL and ADNI is shown in single plots.

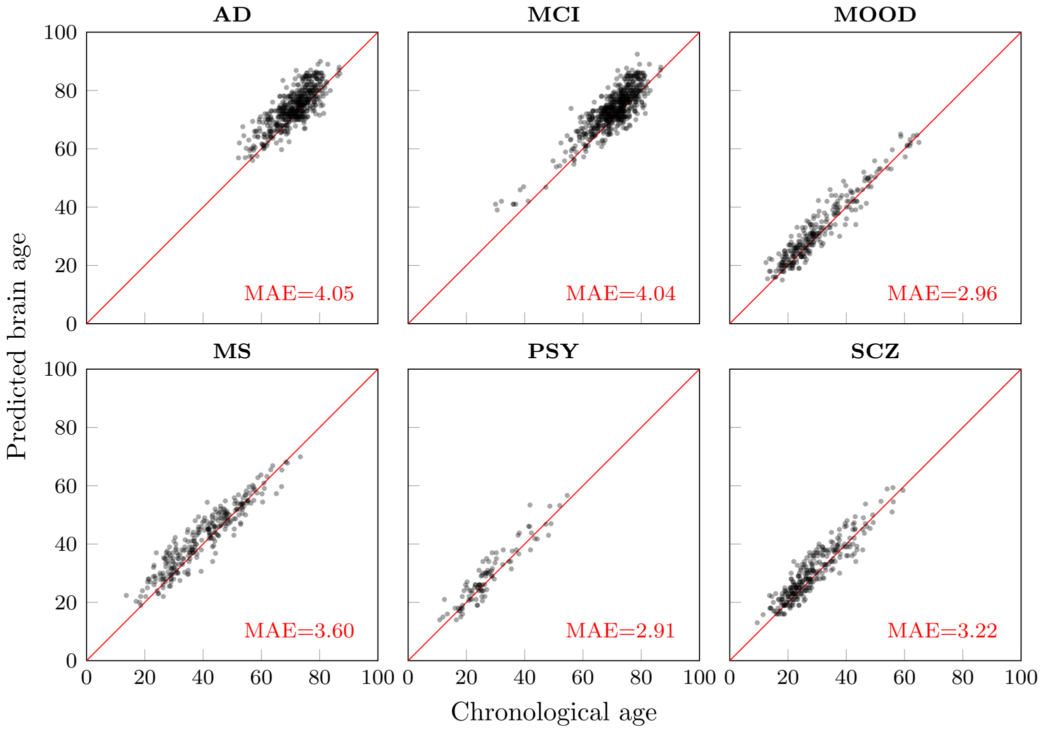


**Supplementary Figure 3: Predictions of SFCN-reg on the control sets used in the clinical classifiers.**

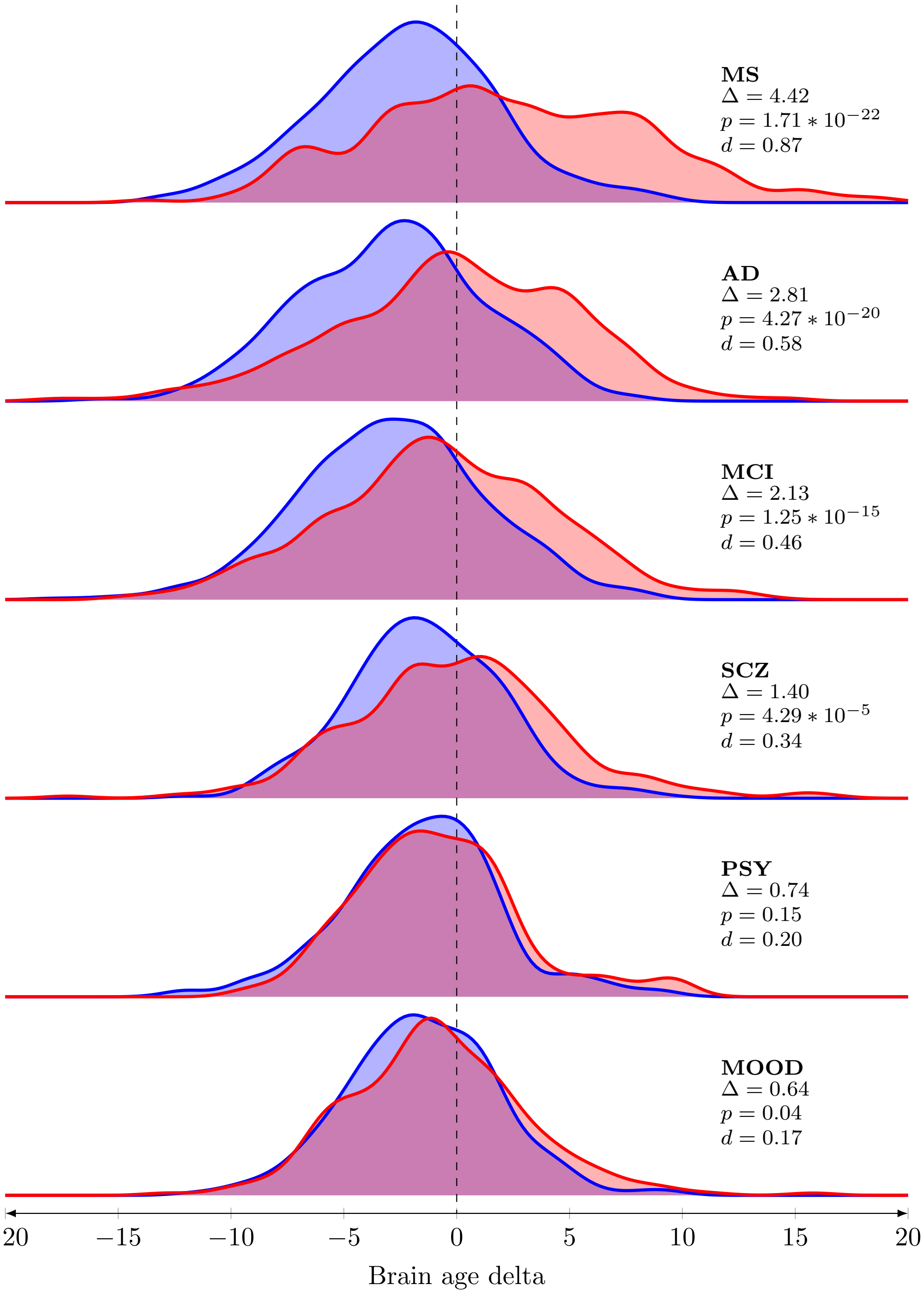


**Supplementary Figure 4: The brain age delta distributions for the individual patient cohorts versus their individually matched control groups.**

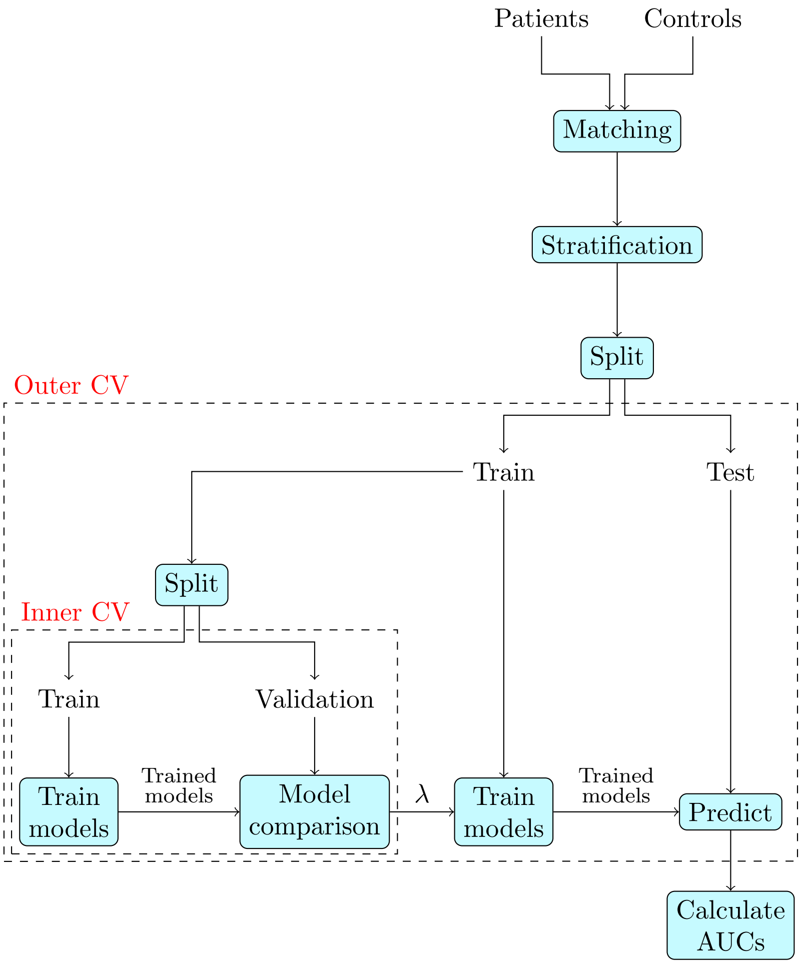


**Supplementary Figure 5: The nested cross-validation (CV) scheme used for the clinical predictors.** The outer CV ensures that the predictions used for the AUC calculations are on unseen data, while the inner loop optimizes the $l_1$-penalty parameter $\lambda$. On each level the data was split into 5 stratified folds, and the same folds were used across the three different LASSO models.

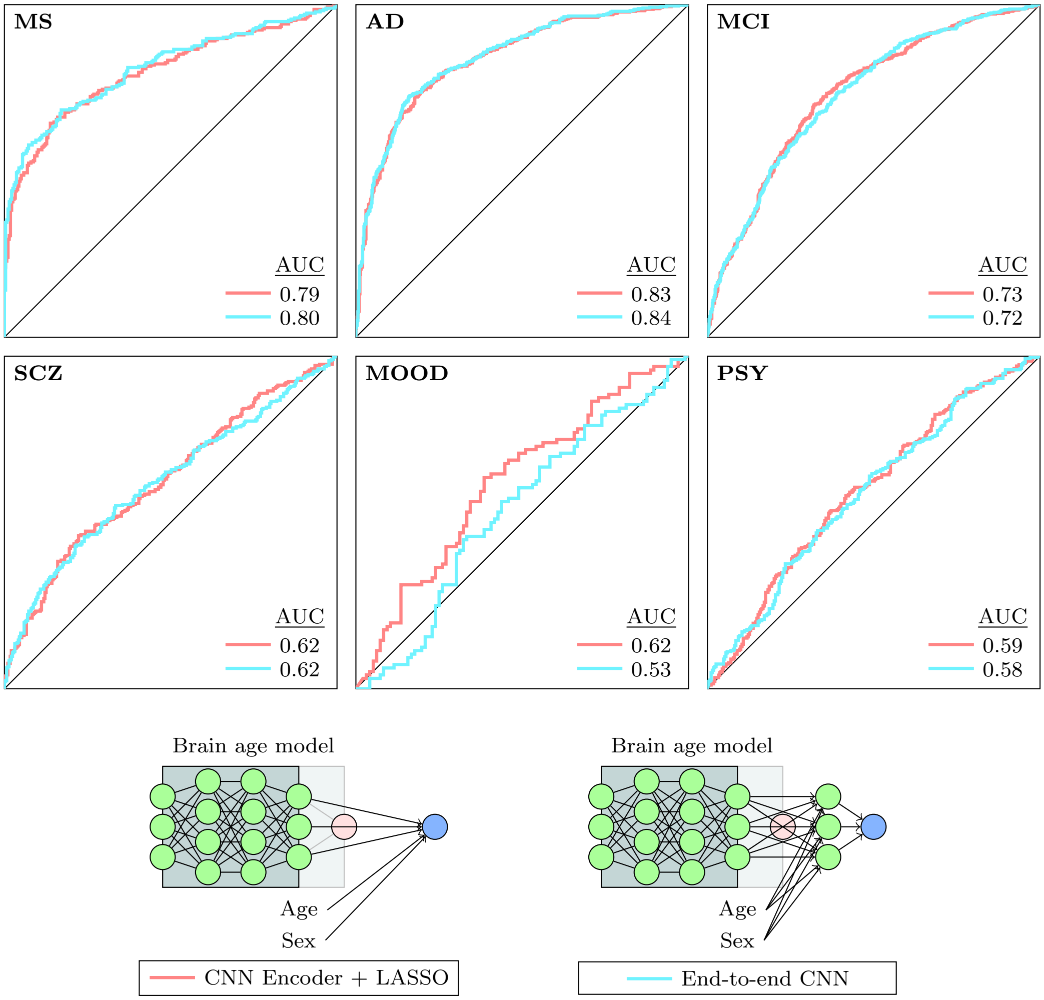


**Supplementary Figure 6: A comparison of the AUCs of the LASSO and CNN clinical predictors.** While both models rely on the features of the brain age model, the LASSO requires a two-step modelling process whereas the End-to-end CNN can be implemented as a single Keras model.

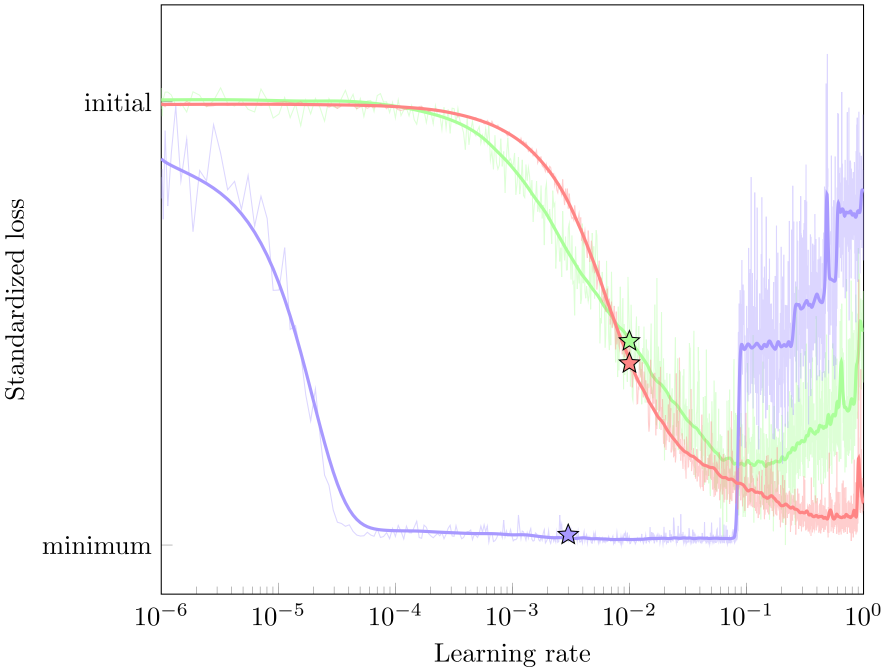


**Supplementary Figure 7: Results of the learning rate sweep for the three model variants.** The sweep investigated the models ability to learn at different learning rates, indicated by a steep descent. The initial learning rates, denoted by the stars, were set manually based on this visualization.

| Name | Size | Subset | Ages | Sex (M/F) | Train | Validation | Test |
|---|---|---|---|---|---|---|---|
| Autism Brain Imaging Data Exchange | 555 | Healthy Controls | 6-56 | 459/96 | 356 | 88 | 111 |
| Autism Brain Imaging Data Exchange II | 184 | Healthy Controls | 8-64 | 114/70 | 118 | 29 | 37 |
| ADHD200 | 606 | Healthy Controls | 7-26 | 313/293 | 388 | 97 | 121 |
| Amsterdam Open MRI Collection | 819 | ID1000 | 19-26 | 390/429 | 524 | 131 | 164 |
| Beijing Normal University Enhanced Sample | 180 | Full | 17-28 | 73/107 | 116 | 28 | 36 |
| Cambridge Center for Aging and Neuroscience | 653 | Full | 18-89 | 323/330 | 418 | 104 | 131 |
| Consortium for Reliability and Reproducibility | 1368 | Full | 6-84 | 672/696 | 880 | 219 | 269 |
| Dallas Lifespan Brain Study | 314 | Full | 21-89 | 117/197 | 201 | 50 | 63 |
| ds000119 | 73 | Full | 8-27 | 30/43 | 47 | 11 | 15 |
| ds000202 | 95 | Full | 18-30 | 0/95 | 61 | 15 | 19 |
| ds000222 | 79 | Full | 21-73 | 38/41 | 51 | 12 | 16 |
| 1000 Functional Connectomes Project | 812 | Full | 8-78 | 377/435 | 520 | 130 | 162 |
| Healthy Brain Network | 1855 | Full | 5-22 | 1183/672 | 1188 | 296 | 371 |
| Human Connectome Project | 1113 | Full | 22-37 | 512/601 | 712 | 178 | 223 |
| Max Planck Institute Leipzig Mind-Brain-Body | 73 | Full | 22-68 | 66/7 | 47 | 11 | 15 |
| Enhanced Nathan Kline Institute - Rockland Sample | 928 | Full | 6-85 | 363/565 | 594 | 148 | 186 |
| Open Access Series of Imaging Studies | 1264 | Healthy Controls | 42-95 | 495/769 | 817 | 188 | 259 |
| Pediactric, Imaging and Neurocognition | 1174 | Full | 3-21 | 610/564 | 752 | 187 | 235 |
| Southwest University Adult Lifespan Dataset | 494 | Full | 19-80 | 187/307 | 316 | 79 | 99 |
| Southwest University Longitudinal Imaging Multimodal | 573 | Full | 17-27 | 254/319 | 367 | 91 | 115 |
| UK Biobank | 40330 | Custom* | 45-82 | 19251/21079 | 25812 | 6452 | 8066 |
| **Total** | **53542** | **-** | **3-95** | **25827/27715** | **34285** | **8544** | **10713** |

**Supplementary Table 1: An overview of the reference dataset.** The dataset was compiled from 21 sources, spanning a wide age range and a multitude of scanners and scanning protocols. How many participants from each source was used for training, validation and testing is indicated in the respective columns.

| Name | Size | Ages | Sex (M/F) |
|---|---|---|---|
| Alzheimer's Disease Neuroimaging Initiative | 416 | 58-96 | 209/207 |
| The Australian Imaging, Behaviour and Lifestyle Flagship Study | 432 | 60-92 | 187/245 |
| IXI | 308 | 19-86 | 129/179 |
| The Norwegian Cognitive NeuroGenetics Sample | 209 | 23-81 | 72/137 |
| StrokeMRI | 341 | 20-94 | 128/213 |
| TOP | 848 | 13-72 | 445/390 |
| **Total** | **2554** | **13-96** | **1170/1371** |

**Supplementary Table 2: An overview of the external dataset used for generalization testing.** The external dataset only contains healthy controls and spans an age range slightly different than that of the reference dataset.

| Cohort | Size | Ages | Source(s) |
|---|---|---|---|
| Alzheimer's disease | 497 | 54-93 | ADNI (338), AIBL (69), Demgen (116) |
| Mild cognitive impairment | 603 | 38-92 | ADNI (402), AIBL (85), Demgen (116) |
| Mixed psychotic diagnoses | 96 | 14-57 | TOP |
| Mood disorders | 269 | 14-65 | TOP |
| Multiple sclerosis | 275 | 19-70 | Multiple sclerosis |
| Schizophrenia | 287 | 13-59 | TOP |

**Supplementary Table 3: An overview of the patient cohorts used in the clinical classifiers.** Each patient cohort has a matched control group, sampled from the same scanners with a resembling age and sex distribution.

| Dataset | Exclusion criteria |
|---|---|
| ABIDE | Patient cohort<br>Timepoints 2+ for longitudinal samples |
| ADHD200 | Patient cohort |
| FCON1000 | Coming from ADHD cohort<br>Beijing Zang cohort removed from test set |
| OASIS3 | Patient cohort<br>Timepoints 2+ for longitudinal samples |
| SLIM | Timepoints 2+ for longitudinal samples |
| UKBB | Having one of the following ICD-10 diagnoses: C70, C71, F[0,99], G[0-99],R[40-47],R90,S06<br>All timepoints for all longitudinal samples |

**Supplementary Table 4: Exclusion criteria for the participants in the reference dataset.**

| Dataset | Source | Comment | Reference(s) |
|---|---|---|---|
| Autism Brain Imaging Dataset Exchange | http://fcon_1000.projects.nitrc.org/ | Primary support for the work by Adriana Di Martino was provided by the NIMH (K23MH087770) and the Leon Levy Foundation. Primary support for the work by Michael P. Milham and the INDI team was provided by gifts from Joseph P. Healy and the Stavros Niarchos Foundation to the Child Mind Institute, as well as by an NIMH award to MPM (R03MH096321). | [1] |
| Autism Brain Imaging Dataset Exchange II | http://fcon_1000.projects.nitrc.org/ | Primary support for the work by Adriana Di Martino and her team was provided by the National Institute of Mental Health (NIMH 5R21MH107045). Primary support for the work by Michael P. Milham and his team provided by the National Institute of Mental Health (NIMH 5R21MH107045); Nathan S. Kline Institute of Psychiatric Research). Additional Support was provided by gifts from Joseph P. Healey, Phyllis Green and Randolph Cowen to the Child Mind Institute. | [2] |
| ADHD200 | http://fcon_1000.projects.nitrc.org/ | F. Xavier Castellanos, David Kennedy, Michael Milham, and Stewart Mostofsky are responsible for the initial conception of the ADHD-200 Consortium. Consortium steering committee includes Jan Buitelaar, F. Xavier Castellanos, Dan Dickstein, Damien Fair, David Kennedy, Beatriz Luna, Michael Milham (Project Coordinator), Stewart Mostofsky, and Julie Schweitzer. Data aggregation and organization was coordinated by the INDI team, which included Saroja Bangaru, David Gutman, Maarten Mennes, and Michael Milham. Web infrastructure and data storage were coordinated by Robert Buccigrossi, Albert Crowley, Christian Hasselgrove, David Kennedy, Kimberly Pohland, and Nina Preuss. The ADHD-200 Global Competition Coordinators were Damien Fair (Chair of Selection Committee, Editor in Chief for Global Competition Special issue) and Michael Milham | [3, 4] |
| Alzheimer's Disease Neuroimaging Initiative | http://adni.loni.usc.edu/ | The ADNI was launched in 2003 as a public-private partnership, led by Principal Investigator Michael W. Weiner, MD. ADNI consists of 4 waves, the later is still ongoing (ADNI 3). A complete listing of ADNI investigators can be found at http://adni.loni.usc.edu/wp-content/uploads/how_to_apply/ADNI_Acknowledgement_List.pdf. Data collection and sharing for this project was funded by the ADNI (NIH Grant U01 AG024904) and DOD ADNI (Department of Defense award number W81XWH-12-2-0012). ADNI is funded by the National Institute on Aging, the National Institute of Biomedical Imaging and Bioengineering, and through generous contributions from the following: AbbVie, Alzheimer's Association; Alzheimer's Drug Discovery Foundation; Araclon Biotech; BioClinica, Inc.; Biogen; Bristol-Myers Squibb Company; CereSpir, Inc.; Cogstate Eisai Inc.; Elan Pharmaceuticals, Inc.; Eli Lilly and Company; EuroImmun; F. Hoffmann-La Roche Ltd and its affiliated company Genentech, Inc.; Fujirebio; GE Healthcare; IXICO Ltd.; Janssen Alzheimer Immunotherapy Research & Development, LLC.; Johnson & Johnson Pharmaceutical Research & Development LLC.; Lumosity; Lundbeck; Merck & Co., Inc.; Meso Scale Diagnostics, LLC.; NeuroRx Research; Neurotrack Technologies; Novartis Pharmaceuticals Corporation; Pfizer Inc.; Piramal Imaging; Servier; Takeda Pharmaceutical Company; and Transition Therapeutics. The Canadian Institutes of Health Research is providing funds to support ADNI clinical sites in Canada and the Saguenay Yout Study. Private sector contributions are facilitated by the Foundation for the National Institutes of Health (http://www.fnih.org). The grantee organization is the Northern California Institute for Research and Education, and the study is coordinated by the Alzheimer's Therapeutic Research Institute at the University of Southern California. ADNI data are disseminated by the Laboratory for Neuro Imaging at the University of Southern California. | |
| The Australian Imaging, Behaviour and Lifestyle Flagship Study | https://aibl.csiro.au/ | Australian Imaging Biomarkers and Lifestyle flagship study of ageing (AIBL) was funded by the Commonwealth Scientific and Industrial Research Organisation (CSIRO), which was made available at the ADNI database (http://www.loni.usc.edu/ADNI). The AIBL researchers contributed data but did not participate in analysis or writing of this report. AIBL researchers are listed at http://www.aibl.csiro.au. Correspondence should be addressed to Christopher Rowe. | |
| The Amsterdam Open MRI Collection | https://nilab-uva.github.io/AOMIC.github.io/ | | [5] |
| Beijing Normal University Enhanced Sample | http://fcon_1000.projects.nitrc.org/ | Financial support for the data used in this project was provided by a grant from the National Natural Science Foundation of China: 30770594 and a grant from the National High Technology Program of China (863): 2008AA02Z405. | [6, 7] |
| Cam-CAN | https://camcan-archive.mrc-cbu.cam.ac.uk/dataaccess/ | Data collection and sharing for this project was provided by the Cambridge Centre for Ageing and Neuroscience (CamCAN). CamCAN funding was provided by the UK Biotechnology and Biological Sciences Research Council (grant number BB/H008217/1), together with support from the UK Medical Research Council and University of Cambridge, UK. | [8, 9] |

| Dataset | Source | Comment | Reference(s) |
|---|---|---|---|
| Consortium for Reliability and Reproducibility | http://fcon_1000.projects.nitrc.org/ | | [10] |
| Demgen | Authors | Supported by the Norwegian National Advisory Unit on Aging and Health | [11, 12] |
| Dallas Lifespan Brain Study | http://fcon_1000.projects.nitrc.org/ | | [13] |
| ds000119 | https://openfmri.org/ | Supported by the National Institutes of Mental Health (NIMH RO1 MH067924). Enami Yasui provided assistance with data collection | [14] |
| ds000202 | https://openfmri.org/ | | [15, 16] |
| ds000222 | https://openfmri.org/ | | [17] |
| 1000 Functional Connectomes Classic Sample | http://fcon_1000.projects.nitrc.org/ | Collected at 33 independent sites by J.J. Pekar, S.H. Mostofsky, S, Colcombe, Y.F. Zang, D. Marguiles, R.L. Buckner, M.J Low, B. Rypma, D.J. Madden, A.C. Evans, S.A.R.B. Rombouts, A. Villringer, S.J. Li, C.Sorg, V. Riedel, B. Biswal, M. Hampson, M.P. Milham, F.X. Castellanos, P. Williamson, M. Hoptman, V.J. Kiviniemi, J. Veijiola, S.M. Smith, C. Mackay, M. Greicius, G. Siegle, K. McMahon, B. Schlaggar, S. Petersen, C.P. Lin, H.S. Mayberg, C.S. Monk, R.D. Seidler, S.J. Peltier | |
| Human Connectome Project | https://www.humanconnectome.org/ | Data were provided [in part] by the Human Connectome Project, MGH-USC Consortium (PIs: Bruce R. Rosen, Arthur W. Toga and Van Wedeen; U01MH093765) funded by the NIH Blueprint Initiative for Neuroscience Research grant; the National Institutes of Health grant P41EB015896; and the Instrumentation Grants S10RR023043, 1S10RR023401, 1S10RR019307. | [18] |
| IXI | http://brain-development.org/ixi-dataset/ | | |
| Max Planck Institut Leipzig Mind-Brain-Body Dataset | http://fcon_1000.projects.nitrc.org/ | | [19] |
| Multiple sclerosis | Authors | The project was supported by grants from The Research Council of Norway (NFR, grant number 240102, 249795, and 223273) and the South-Eastern Health Authorities of Norway (grant number 257955, 2014097). | [20] |
| The Norwegian Cognitive NeuroGenetics Sample (NCNG) | Authors | Supported by grants from the the Research Council of Norway (177458/V50 and 231286/F20). | [21] |
| Enhanced Nathan Kline Institute - Rockland Sample | http://fcon_1000.projects.nitrc.org/ | Principal support for the enhanced NKI-RS project is provided by the NIMH BRAINS R01MH094639-01 (PI Milham). Funding for key personnel also provided in part by the New York State Office of Mental Health and Research Foundation for Mental Hygiene. Funding for the decompression and augmentation of administrative and phenotypic protocols provided by a grant from the Child Mind Institute (1FDN2012-1). Additional personnel support provided by the Center for the Developing Brain at the Child Mind Institute, as well as NIMH R01MH081218, R01MH083246, and R21MH084126. Project support also provided by the NKI Center for Advanced Brain Imaging (CABI), the Brain Research Foundation, and the Stavros Niarchos Foundation. | [22] |
| Open Access Series of Imaging Studies 3 | http://www.oasis-brains.org/ | Supported by grants P50 AG05681, P01 AG03991, R01 AG021910, P50 MH071616, U24 RR021382, R01 MH56584. | [23, 24] |
| Pediatric Imaging, Neurocognition and Genetics | http://pingstudy.ucsd.edu/ | Data used in the preparation of this article were obtained from the Pediatric Imaging, Neurocognition and Genetics (PING) Study database (www.chd.ucsd.edu/research/ping-study.html, now shared through the NIMH Data Archive (NDA)). PING was a multisite, cross-sectional study that recruited more than 1,700 participants aged 3 to 20 years. The study was supported by award number RC2DA029475 from the National Institute on Drug Abuse with additional support for data sharing provided by the Eunice Kennedy Shriver National Institute of Child Health & Human Development under award number R01HD061414. A list of participating sites and study investigators can be found at https://ping-dataportal.ucsd.edu/sharing/Authors10222012.pdf. PING investigators designed and implemented the study and/or provided data but did not necessarily participate in analysis or writing of this report. This publication is solely the responsibility of the authors and does not necessarily represent the views of the National Institutes of Health or PING investigators. | [25] |

| Dataset | Source | Comment | Reference(s) |
|---|---|---|---|
| Southwest University Adult Lifespan Dataset | http://fcon_1000.projects.nitrc.org/ | | [26] |
| Southwest University Longitudinal Imaging Multimodal Brain Data | http://fcon_1000.projects.nitrc.org/ | Support was provided by grant numbers 31271087; 31470981; 31571137, 31500885, SWU1509383, SWU1509451, cstc2015jcyjA10106, 151023, 2015M572423, 2015M580767, Xm2015037, 14JJD880009. | [27, 28] |
| StrokeMRI | Authors | Supported by the Research Council of Norway (249795, 248238), the South-Eastern Norway Regional Health Authority (2014097, 2015044, 2015073, 2016083), and the Norwegian ExtraFoundation for Health and Rehabilitation (2015/FO5146) | [29] |
| TOP | Authors | Supported by several grants from the Research Council of Norway, and the South-Eastern Norway Regional Health Authority | [30–32] |
| UK BioBank | https://www.ukbiobank.ac.uk/ | This research has been conducted using the UK Biobank Resource (access code 27412). | [33, 34] |

**Supplementary Table 5: An overview of the sources of all the datasets used in the study.**

| Dataset | Number of scanners/protocols | Parameters |
|---|---|---|
| ABIDE | 20 | http://fcon_1000.projects.nitrc.org/indi/abide/scan_params/ |
| ABIDE II | 16 | http://fcon_1000.projects.nitrc.org/indi/abide/scan_params/ |
| ADHD200 | 6 | Philips 1.5 T Gyroscan: TR=8ms, TE=3.76ms, FA=8°<br>Siemens 3T Allegra: TR=2530ms, TE=3.25ms, FA=8°<br>Siemens 3T Trio: TR=2300ms, TE=3.58ms, FA=10°<br>Siemens 3T Trio: TR=1700ms, TE=3.92ms, FA=12°<br>Siemens 3T Trio: TR=2100ms, TE=3.43ms, FA=8°<br>Siemens 3T Trio: TR=2400ms, TE=3.08ms, FA=8° |
| ADNI | 57 | http://adni.loni.usc.edu/methods/mri-tool/mri-analysis/ |
| AIBL | 3 | Siemens 1.5T Avanto: 3D MP-RAGE, TR = 2300ms, TE = 2.98ms, TI = 900, FA = 9°<br>Siemens 3.0T Verio: 3D MP-RAGE, TR = 2300ms, TE = 2.98ms, TI = 900, FA = 9°<br>Siemens 3.0T Tim Trio Siemens: 3D MP-RAGE, TR = 2300ms, TE = 2.98ms, TI = 900, FA = 9° |
| AOMIC | 1 | Philips 3T Intera: 3D MPRAGE TR=8.1ms, TE=3.7ms, FA=8° |
| Beijing Enhanced | 1 | SIEMENS TRIO 3T: MPRAGE,TR=2530ms,TE=3.39ms,FA=7° |
| Cam-CAN | 1 | Siemens 3T Trio: TR=2250ms, TE=2.99ms, FA=9° |
| CoRR | 34 | http://fcon_1000.projects.nitrc.org/indi/CoRR/html/_static/scan_parameters/ |
| Demgen | 2 | GE 3T Signa HDxT: TR=7.8ms, TE=2.956ms, FA=12°<br>GE 3T Discovery GE750: TR=8.16ms, TE=3.18ms, FA=12° |
| DLBS | 1 | Philips 3T: TR=8.135ms, TE=3.7ms, FA=18° |
| ds000119 | 1 | Siemens 3T Allegra: TR=1570ms, TE=3.04ms, FA=8° |
| ds00202 | 1 | Philips 3T Achieva: TR=7.6ms, TE=3.7ms,, FA=8° |
| ds000222 | 1 | Siemens 3T Trio: TR=1550ms, TE=2.34ms, FA=9° |
| FCON 1000 | 28 | http://fcon_1000.projects.nitrc.org/fcpClassic/FcpTable.html |
| HCP | 1 | Customized 3T scanner: TR=2400ms, TE=2.14, FA=8° |
| IXI | 3 | Philips 3T: TR=9.6ms, TE=4.6ms, FA=8°<br>Philips 1.5T: TR=9.8ms, TE=4.6ms, FA=8°<br>GE 1.5T: TR=6.0ms, TE=2.5ms |
| MPI-LEMON | 1 | Siemens MAGNETOM Verio 3T: MPRAGE,TR=680ms, TE1=5.19ms,TE2=7.65ms,FA=60° |
| Multiple sclerosis | 2 | Siements 1.5T Avanto: TR=2400ms, TE=3.61ms, FA=8°<br>GE 3T Discovery GE750: TR=8.16ms, TE=3.18ms, FA=12° |
| NCNG | 1 | Siements 1.5T Avanto: TR=2400ms, TE=3.61ms, FA=8° |
| NKI Rockland | 2 | Siemens MAGNETOM TrioTim 3T: TR=1900ms, TE=2.52ms, FA=9°<br>Siemens MAGNETOM TrioTim 3T: TR=2600ms, TE=3.02ms, FA=8° |
| OASIS 3 | 1 | Siemens 1.5T Vision: TR=9.7ms, TE=4ms, FA=10° |
| PING | 11 | http://pingstudy.ucsd.edu/resources/neuroimaging-cores.html |
| SALD | 1 | Siemens 3T Trio: TR=1900ms, TE=2.52ms, FA=9° |
| SLIM | 1 | Siemens 3T Trio: TR=1900ms, TE=2.52ms, FA=9° |
| StrokeMRI | 2 | GE 3T Signa HDxT: TR=7.8ms, TE=2.956ms, FA=12°<br>GE 3T Discovery GE750: TR=8.16ms, TE=3.18ms, FA=12° |
| TOP | 4 | Siemens 1.5T Sonata: TR=2730ms, TE=3.93ms, FA=7°<br>GE 3T Signa HDxT: TR=7.8ms, TE=2.956ms, FA=12°<br>(one subset with HNS coil, one subset with 8HRBRAIN coil)<br>GE 3T Discovery GE750: TR=8.16ms, TE=3.18ms, FA=12° |
| UK Biobank | 3 | Siemens 3T Skyra: TR=2000ms, TE=2.01ms, FA=8° (3 identical scanning sites) |

**Supplementary Table 6: An overview of the scanning protocols used in the datasets.**

| Category | Columns |
|---|---|
| Alcohol | 'Alcohol drinker status', 'Alcohol intake frequency', 'Former alcohol drinker', 'Average monthly red wine intake', 'Average monthly champagne plus white wine intake', 'Average monthly beer plus cider intake', 'Average monthly spirits intake', 'Average monthly fortified wine intake', 'Average monthly intake of other alcoholic drinks', 'Average weekly red wine intake', 'Average weekly champagne plus white wine intake', 'Average weekly beer plus cider intake', 'Average weekly spirits intake', 'Average weekly fortified wine intake', 'Average weekly intake of other alcoholic drinks', 'Alcohol usually taken with meals', 'Alcohol intake versus 10 years previously', 'Reason for reducing amount of alcohol drunk', 'Reason former drinker stopped drinking alcohol' |
| Biochemical | 'Basophill count', 'Basophill percentage', 'Eosinophill count', 'Eosinophill percentage', 'Haematocrit percentage', 'Haemoglobin concentration', 'High light scatter reticulocyte count', 'High light scatter reticulocyte percentage', 'Immature reticulocyte fraction', 'Lymphocyte count', 'Lymphocyte percentage', 'Mean corpuscular haemoglobin', 'Mean corpuscular haemoglobin concentration', 'Mean corpuscular volume', 'Mean platelet (thrombocyte) volume', 'Mean reticulocyte volume', 'Mean sphered cell volume', 'Monocyte count', 'Monocyte percentage', 'Neutrophill count', 'Neutrophill percentage', 'Nucleated red blood cell count', 'Nucleated red blood cell percentage', 'Platelet count', 'Platelet crit', 'Platelet distribution width', 'Red blood cell (erythrocyte) count', 'Red blood cell (erythrocyte) distribution width', 'Reticulocyte count', 'Reticulocyte percentage', 'White blood cell (leukocyte) count', 'Alanine aminotransferase', 'Albumin', 'Alkaline phosphatase', 'Apolipoprotein A', 'Apolipoprotein B', 'Aspartate aminotransferase', 'C-reactive protein', 'Calcium', 'Cholesterol', 'Creatinine', 'Cystatin C', 'Direct bilirubin', 'Gamma glutamyltransferase', 'Glucose', 'Glycated haemoglobin (HbA1c)', 'HDL cholesterol', 'IGF-1', 'LDL direct', 'Lipoprotein A', 'Oestradiol', 'Phosphate', 'Rheumatoid factor', 'SHBG', 'Testosterone', 'Total bilirubin', 'Total protein', 'Triglycerides', 'Urate', 'Urea', 'Vitamin D', '3-Hydroxybutyrate', 'Acetate', 'Acetoacetate', 'Acetone', 'Alanine', 'Albumin', 'Apolipoprotein A1', 'Apolipoprotein B', 'Average Diameter for HDL Particles', 'Average Diameter for LDL Particles', 'Average Diameter for VLDL Particles', 'Cholesterol in Chylomicrons and Extremely Large VLDL', 'Cholesterol in IDL', 'Cholesterol in Large HDL', 'Cholesterol in Large LDL', 'Cholesterol in Large VLDL', 'Cholesterol in Medium HDL', 'Cholesterol in Medium LDL', 'Cholesterol in Medium VLDL', 'Cholesterol in Small HDL', 'Cholesterol in Small LDL', 'Cholesterol in Small VLDL', 'Cholesterol in Very Large HDL', 'Cholesterol in Very Large VLDL', 'Cholesterol in Very Small VLDL', 'Cholesteryl Esters in Chylomicrons and Extremely Large VLDL', 'Cholesteryl Esters in HDL', 'Cholesteryl Esters in IDL', 'Cholesteryl Esters in LDL', 'Cholesteryl Esters in Large HDL', 'Cholesteryl Esters in Large LDL', 'Cholesteryl Esters in Large VLDL', 'Cholesteryl Esters in Medium HDL', 'Cholesteryl Esters in Medium LDL', 'Cholesteryl Esters in Medium VLDL', 'Cholesteryl Esters in Small HDL', 'Cholesteryl Esters in Small LDL', 'Cholesteryl Esters in Small VLDL', 'Cholesteryl Esters in VLDL', 'Cholesteryl Esters in Very Large HDL', 'Cholesteryl Esters in Very Large VLDL', 'Cholesteryl Esters in Very Small VLDL', 'Citrate', 'Clinical LDL Cholesterol', 'Concentration of Chylomicrons and Extremely Large VLDL Particles', 'Concentration of HDL Particles', 'Concentration of IDL Particles', 'Concentration of LDL Particles', 'Concentration of Large HDL Particles', 'Concentration of Large LDL Particles', 'Concentration of Large VLDL Particles', 'Concentration of Medium HDL Particles', 'Concentration of Medium LDL Particles', 'Concentration of Medium VLDL Particles', 'Concentration of Small HDL Particles', 'Concentration of Small LDL Particles', 'Concentration of Small VLDL Particles', 'Concentration of VLDL Particles', 'Concentration of Very Large HDL Particles', 'Concentration of Very Large VLDL Particles', 'Concentration of Very Small VLDL Particles', 'Creatinine', 'Degree of Unsaturation', 'Docosahexaenoic Acid', 'Free Cholesterol in Chylomicrons and Extremely Large VLDL', 'Free Cholesterol in HDL', 'Free Cholesterol in IDL', 'Free Cholesterol in LDL', 'Free Cholesterol in Large HDL', 'Free Cholesterol in Large LDL', 'Free Cholesterol in Large VLDL', 'Free Cholesterol in Medium HDL', 'Free Cholesterol in Medium LDL', 'Free Cholesterol in Medium VLDL', 'Free Cholesterol in Small HDL', 'Free Cholesterol in Small LDL', 'Free Cholesterol in Small VLDL', 'Free Cholesterol in VLDL', 'Free Cholesterol in Very Large HDL', 'Free Cholesterol in Very Large VLDL', 'Free Cholesterol in Very Small VLDL', 'Glucose', 'Glutamine', 'Glycine', 'Glycoprotein Acetyls', 'HDL Cholesterol', 'Histidine', 'Isoleucine', 'LDL Cholesterol', 'Lactate', 'Leucine', 'Linoleic Acid', 'Monounsaturated Fatty Acids', 'Omega-3 Fatty Acids', 'Omega-6 Fatty Acids', 'Phenylalanine', 'Phosphatidylcholines', 'Phosphoglycerides', 'Phospholipids in Chylomicrons and Extremely Large VLDL', 'Phospholipids in HDL', 'Phospholipids in IDL', 'Phospholipids in LDL', 'Phospholipids in Large HDL', 'Phospholipids in Large LDL', 'Phospholipids in Large VLDL', 'Phospholipids in Medium HDL', 'Phospholipids in Medium LDL', 'Phospholipids in Medium VLDL', 'Phospholipids in Small HDL', 'Phospholipids in Small LDL', 'Phospholipids in Small VLDL', 'Phospholipids in VLDL', 'Phospholipids in Very Large HDL', 'Phospholipids in Very Large VLDL', 'Phospholipids in Very Small VLDL', 'Polyunsaturated Fatty Acids', 'Pyruvate', 'Remnant Cholesterol (Non-HDL, Non-LDL -Cholesterol)', 'Saturated Fatty Acids', 'Sphingomyelins', 'Total Cholesterol', 'Total Cholesterol Minus HDL-C', 'Total Cholines', 'Total Concentration of Branched-Chain Amino Acids (Leucine + Isoleucine + Valine)', 'Total Concentration of Lipoprotein Particles', 'Total Esterified Cholesterol', 'Total Fatty Acids', 'Total Free Cholesterol', 'Total Lipids in Chylomicrons and Extremely Large VLDL', 'Total Lipids in HDL', 'Total Lipids in IDL', 'Total Lipids in LDL', 'Total Lipids in Large HDL', 'Total Lipids in Large LDL', 'Total Lipids in Large VLDL', 'Total Lipids in Lipoprotein Particles', 'Total Lipids in Medium HDL', 'Total Lipids in Medium LDL', 'Total Lipids in Medium VLDL', 'Total Lipids in Small HDL', 'Total Lipids in Small LDL', 'Total Lipids in Small VLDL', 'Total Lipids in VLDL', 'Total Lipids in Very Large HDL', 'Total Lipids in Very Large VLDL', 'Total Lipids in Very Small VLDL', 'Total Phospholipids in Lipoprotein Particles', 'Total Triglycerides', 'Triglycerides in Chylomicrons and Extremely Large VLDL', 'Triglycerides in HDL', 'Triglycerides in IDL', 'Triglycerides in LDL', 'Triglycerides in Large HDL', 'Triglycerides in Large LDL', 'Triglycerides in Large VLDL', 'Triglycerides in Medium HDL', 'Triglycerides in Medium LDL', 'Triglycerides in Medium VLDL', 'Triglycerides in Small HDL', 'Triglycerides in Small LDL', 'Triglycerides in Small VLDL', 'Triglycerides in VLDL', 'Triglycerides in Very Large HDL', 'Triglycerides in Very Large VLDL', 'Triglycerides in Very Small VLDL', 'Tyrosine', 'VLDL Cholesterol', 'Valine', 'Creatinine (enzymatic) in urine', 'Microalbumin in urine', 'Potassium in urine', 'Sodium in urine' |

| | |
|---|---|
| Cognitive | 'Mean time to correctly identify matches"', 'Digits entered correctly', 'Maximum digits remembered correctly', 'Time first key touched', 'Time last key touched', 'Time elapsed', 'Time to complete test', 'Fluid intelligence score', 'Number of fluid intelligence questions attempted within time limit', 'Attempted fluid intelligence (FI) test', 'FI1 : numeric addition test', 'FI2 : identify largest number', 'FI3 : word interpolation', 'FI4 : positional arithmetic', 'FI5 : family relationship calculation', 'FI6 : conditional arithmetic', 'FI7 : synonym', 'FI8 : chained arithmetic', 'FI9 : concept interpolation', 'FI10 : arithmetic sequence recognition', 'FI11 : antonym', 'FI12 : square sequence recognition', 'FI13 : subset inclusion logic', 'Duration to complete numeric path (trail #1)', 'Duration to complete alphanumeric path (trail #2)', 'Total errors traversing numeric path (trail #1)', 'Total errors traversing alphanumeric path (trail #2)', 'Interval between previous point and current one in numeric path (trail #1)', 'Interval between previous point and current one in alphanumeric path (trail #2)', 'Errors before selecting correct item in numeric path (trail #1)', 'Errors before selecting correct item in alphanumeric path (trail #2)', 'Number of puzzles correctly solved', 'Number of puzzles viewed', 'Duration spent answering each puzzle', 'Vocabulary level', 'Number of word pairs correctly associated', 'Word associated with huge', 'Word associated with happy', 'Word associated with tattered', 'Word associated with old', 'Word associated with long', 'Word associated with red', 'Word associated with sulking', 'Word associated with pretty', 'Word associated with tiny', 'Word associated with new', 'Prospective memory result', 'Number of incorrect matches in round', 'Time to complete round', 'Number of correct matches in round' |
| Diet | 'Cooked vegetable intake', 'Salad / raw vegetable intake', 'Fresh fruit intake', 'Dried fruit intake', 'Oily fish intake', 'Non-oily fish intake', 'Processed meat intake', 'Poultry intake', 'Beef intake', 'Lamb/mutton intake', 'Pork intake', 'Age when last ate meat', 'Never eat eggs, dairy, wheat, sugar', 'Cheese intake', 'Milk type used', 'Spread type', 'Bread intake', 'Cereal intake', 'Salt added to food', 'Tea intake', 'Coffee intake', 'Water intake', 'Major dietary changes in the last 5 years', 'Variation in diet' |
| Early life factors | 'Ease of skin tanning', 'Hair colour (natural, before greying)', 'Country of birth', 'Breastfed as a baby', 'Comparative body size at age 10', 'Comparative height size at age 10', 'Handedness', 'Adopted as a child', 'Part of a multiple birth', 'Relative age of first facial hair', 'Relative age voice broke', 'Birth weight', 'Country of Birth (non-UK origin)' |
| Family | 'Illnesses of adopted father', 'Illnesses of adopted mother', 'Illnesses of adopted siblings', 'Illnesses of father', 'Illnesses of mother', 'Illnesses of siblings', 'Father still alive', "Father's age", "Father's age at death", 'Mother still alive', "Mother's age", "Mother's age at death", 'Number of full brothers', 'Number of adopted brothers', 'Number of full sisters', 'Number of adopted sisters', 'Number of older siblings', 'Non-accidental death in close genetic family' |
| Health related | 'Facial ageing', 'Mouth/teeth dental problems', 'Overall health rating', 'Long-standing illness, disability or infirmity', 'Falls in the last year', 'Wheeze or whistling in the chest in last year', 'Shortness of breath walking on level ground', 'Pain type(s) experienced in last month', 'Headaches for 3+ months', 'Facial pains for 3+ months', 'Neck/shoulder pain for 3+ months', 'Back pain for 3+ months', 'Stomach/abdominal pain for 3+ months', 'General pain for 3+ months', 'Had major operations', 'Had other major operations', 'Vascular/heart problems diagnosed by doctor', 'Age heart attack diagnosed', 'Age angina diagnosed', 'Age stroke diagnosed', 'Age high blood pressure diagnosed', 'Blood clot, DVT, bronchitis, emphysema, asthma, rhinitis, eczema, allergy diagnosed by doctor', 'Age deep-vein thrombosis (DVT, blood clot in leg) diagnosed', 'Age pulmonary embolism (blood clot in lung) diagnosed', 'Age emphysema/chronic bronchitis diagnosed', 'Age asthma diagnosed', 'Age hay fever, rhinitis or eczema diagnosed', 'Diabetes diagnosed by doctor', 'Gestational diabetes only', 'Age diabetes diagnosed', 'Started insulin within one year diagnosis of diabetes', 'Cancer diagnosed by doctor', 'Fractured/broken bones in last 5 years', 'Fractured bone site(s)', 'Fracture resulting from simple fall', 'Other serious medical condition/disability diagnosed by doctor', 'Medication for cholesterol, blood pressure or diabetes', 'Medication for cholesterol, blood pressure, diabetes, or take exogenous hormones', 'Taking other prescription medications', 'Medication for pain relief, constipation, heartburn', 'Vitamin and mineral supplements', 'Mineral and other dietary supplements', 'Hearing difficulty/problems', 'Hearing aid user', 'Cochlear implant', 'Tinnitus', 'Hair/balding pattern', 'Age when periods started (menarche)', 'Had menopause', 'Age at menopause (last menstrual period)', 'Time since last menstrual period', 'Length of menstrual cycle', 'Menstruating today', 'Number of live births', 'Age at first live birth', 'Age at last live birth', 'Ever had stillbirth, spontaneous miscarriage or termination', 'Ever taken oral contraceptive pill', 'Age started oral contraceptive pill', 'Ever had hysterectomy (womb removed)', 'Cancer code, self-reported', 'Cancer year/age first occurred', 'Pregnant', 'Pace-maker', 'Glasses worn/required (left)', 'Glasses worn/required (right)', 'Wears glasses or contact lenses', 'Age started wearing glasses or contact lenses', 'Reason for glasses/contact lenses', 'Other eye problems', 'Eye problems/disorders', 'Age when diabetes-related eye disease diagnosed', 'Age glaucoma diagnosed', 'Age when loss of vision due to injury or trauma diagnosed', 'Age cataract diagnosed', 'Age macular degeneration diagnosed', 'Age other serious eye condition diagnosed' |
| Lifestyle | 'Length of mobile phone use', 'Weekly use of mobile phone in last 3 months', 'Plays computer games', 'Sleep duration', 'Getting up in morning', 'Morning/evening person', 'Nap during day', 'Sleeplessness/insomnia', 'Daytime dozing/sleeping (narcolepsy)', 'Time spend outdoors in summer', 'Time spent outdoors in winter', 'Use of sun/uv protection', 'Frequency of solarium/sunlamp use', 'Age first had sexual intercourse', 'Lifetime number of sexual partners', 'Ever had same-sex intercourse', 'Lifetime number of same-sex sexual partners', 'Frequency of friend/family visits', 'Leisure/social activities' |
| Mental health | 'Able to confide', 'Bipolar and major depression status', 'Bipolar disorder status', 'Neuroticism score', 'Probable recurrent major depression (moderate)', 'Probable recurrent major depression (severe)', 'Single episode of probable major depression', 'Mood swings', 'Miserableness', 'Irritability', 'Sensitivity / hurt feelings', 'Fed-up feelings', 'Nervous feelings', 'Worrier / anxious feelings', "Tense / 'highly strung'", 'Worry too long after embarrassment', "Suffer from 'nerves'", 'Loneliness, isolation', 'Guilty feelings', 'Risk taking', 'Happiness', 'Work/job satisfaction', 'Health satisfaction', 'Family relationship satisfaction', 'Friendships satisfaction', 'Financial situation satisfaction', 'Seen doctor (GP) for nerves, anxiety, tension or depression', 'Seen a psychiatrist for nerves, anxiety, tension or depression' |

| | |
|---|---|
| | 'Ever depressed for a whole week', 'Longest period of depression', 'Number of depression episodes', 'Ever unenthusiastic/disinterested for a whole week', 'Longest period of unenthusiasm / disinterest', 'Number of unenthusiastic/disinterested episodes', 'Ever manic/hyper for 2 days', 'Ever highly irritable/argumentative for 2 days', 'Manic/hyper symptoms', 'Length of longest manic/irritable episode', 'Severity of manic/irritable episodes', 'Illness, injury, bereavement, stress in last 2 years' |
| Microbial infection | 'HSV-1 seropositivity for Herpes Simplex virus-1', 'HSV-2 seropositivity for Herpes Simplex virus-2', 'VZV seropositivity for Varicella Zoster Virus', 'EBV seropositivity for Epstein-Barr Virus', 'CMV seropositivity for Human Cytomegalovirus', 'HHV-6 overall seropositivity for Human Herpesvirus-6', 'HHV-6A seropositivity for Human Herpesvirus-6', 'HHV-6B seropositivity for Human Herpesvirus-6', 'HHV-7 seropositivity for Human Herpesvirus-7', "KSHV seropositivity for Kaposi's Sarcoma-Associated Herpesvirus", 'HBV seropositivity for Hepatitis B Virus', 'HCV seropositivity for Hepatitis C Virus', 'T. gondii seropositivity for Toxoplasma gondii', 'HTLV-1 seropositivity for Human T-Lymphotropic Virus 1', 'HIV-1 seropositivity for Human Immunodeficiency Virus', 'BKV seropositivity for Human Polyomavirus BKV', 'JCV seropositivity for Human Polyomavirus JCV', 'MCV seropositivity for Merkel Cell Polyomavirus', 'HPV 16 Definition I seropositivity for Human Papillomavirus type-16', 'HPV 16 Definition II seropositivity for Human Papillomavirus type-16', 'HPV 18 seropositivity for Human Papillomavirus type-18', 'C. trachomatis Definition I seropositivity for Chlamydia trachomatis', 'C. trachomatis Definition II seropositivity for Chlamydia trachomatis', 'H. pylori Definition I seropositivity for Helicobacter pylori', 'H. pylori Definition II seropositivity for Helicobacter pylori' |
| Physical measures | 'Diastolic blood pressure, automated reading', 'Diastolic blood pressure, manual reading', 'Pulse rate (during blood-pressure measurement)', 'Pulse rate, automated reading', 'Systolic blood pressure, automated reading', 'Systolic blood pressure, manual reading', 'Absence of notch position in the pulse waveform', 'Arterial pulse-wave stiffness device ID', 'Arterial stiffness device ID', 'Position of pulse wave notch', 'Position of the pulse wave peak', 'Position of the shoulder on the pulse waveform', 'Pulse rate', 'Pulse wave Arterial Stiffness index', 'Pulse wave peak to peak time', 'Pulse wave reflection index', 'Pulse wave velocity (manual entry)', 'Hand grip strength (left)', 'Hand grip strength (right)', 'Height', 'Weight (pre-imaging)', 'Waist circumference', 'Weight', 'Body mass index (BMI)', 'Hip circumference', 'Basal metabolic rate', 'Body fat percentage', 'Ventricular rate' |
| Smoking | 'Ever smoked', 'Pack years adult smoking as proportion of life span exposed to smoking', 'Ever smoked', 'Pack years adult smoking as proportion of life span exposed to smoking', 'Pack years of smoking', 'Smoking status', 'Current tobacco smoking', 'Past tobacco smoking', 'Light smokers, at least 100 smokes in lifetime', 'Age started smoking in current smokers', 'Type of tobacco currently smoked', 'Previously smoked cigarettes on most/all days', 'Number of cigarettes currently smoked daily (current cigarette smokers)', 'Age stopped smoking cigarettes (current cigar/pipe or previous cigarette smoker)', 'Number of cigarettes previously smoked daily (current cigar/pipe smokers)', 'Time from waking to first cigarette', 'Difficulty not smoking for 1 day', 'Ever tried to stop smoking', 'Wants to stop smoking', 'Smoking compared to 10 years previous', 'Age started smoking in former smokers', 'Type of tobacco previously smoked', 'Number of cigarettes previously smoked daily', 'Age stopped smoking', 'Ever stopped smoking for 6+ months', 'Number of unsuccessful stop-smoking attempts', 'Likelihood of resuming smoking', 'Smoking/smokers in household', 'Exposure to tobacco smoke at home', 'Exposure to tobacco smoke outside home', 'Maternal smoking around birth' |
| Socioeconomic | 'Number in household', 'Number of vehicles in household', 'Average total household income before tax', 'Current employment status', 'Job involves mainly walking or standing', 'Job involves heavy manual or physical work', 'Qualifications (Education)', 'Age completed full time education', 'Attendance/disability/mobility allowance', 'Private healthcare' |

**Supplementary Table 7: The columns used in the PheWAS, manually divided into the thirteen categories.**

| Loss function | Dropout | Weight decay | $MAE_{TRAIN}$ | $MAE_{VAL}$ |
|---|---|---|---|---|
| Soft Classification | 0.5 | 1e-3 | 1.79 | 2.34 |
| **Soft Classification** | **0.3** | **1e-3** | **1.57** | **2.28** |
| Soft Classification | 0.3 | 1e-4 | 1.99 | 2.42 |
| Regression | 0.3 | 1e-3 | 1.96 | 2.67 |
| **Regression** | **0.2** | **1e-4** | **1.67** | **2.51** |
| Regression | 0.0 | 0.0 | 1.26 | 2.53 |
| **Ranking** | **0.3** | **1e-3** | **2.38** | **2.59** |
| Ranking | 0.2 | 1e-4 | 2.95 | 3.03 |
| Ranking | 0.5 | 1e-3 | 2.53 | 2.70 |

**Supplementary Table 8: The hyperparameter settings tested for each of the three model variants.** The best version for each model variant was selected based on the Mean Absolute Error in the validation set ($MAE_{VAL}$). The hyperparameter settings were determined sequentially, based on the performance of the previous runs.

| Model | $\mathrm{MAE}_{\mathrm{TEST}}$ | $\mathrm{MAE}_{\mathrm{EXTERNAL}}$ | Generalization gap $\left(\frac{MAE_{EXTERNAL}}{MAE_{TEST}}\right)$ |
|---|---|---|---|
| SFCN-sm | **2.23** | 5.04 | 2.26 |
| SFCN-reg | 2.47 | **3.90** | **1.57** |
| SFCN-rank | 2.55 | 5.92 | 2.32 |

**Supplementary Table 9: Results of the final model comparison.** The test Mean Absolute Error ($\mathrm{MAE}_{\mathrm{TEST}}$) was computed from the test set, a portion of the reference dataset unseen during optimization. The external Mean Absolute Error ($\mathrm{MAE}_{\mathrm{EXTERNAL}}$) was computed on the external dataset, comprised of different origins, containing other scanners and a diverging age distribution compared to that of the training set. The generalization gap indicates the expected degree of deterioriation when transferring to this new dataset.
